# Dysfunction of proprioceptive sensory synapses is a pathogenic event and therapeutic target in mice and humans with spinal muscular atrophy

**DOI:** 10.1101/2024.06.03.24308132

**Authors:** CM Simon, N Delestree, J Montes, F Gerstner, E Carranza, L Sowoidnich, JM Buettner, JG Pagiazitis, G Prat-Ortega, S Ensel, S Donadio, JL Garcia, P Kratimenos, WK Chung, CJ Sumner, LH Weimer, E Pirondini, M Capogrosso, L Pellizzoni, DC De Vivo, GZ Mentis

## Abstract

Spinal muscular atrophy (SMA) is a neurodegenerative disease characterized by a varying degree of severity that correlates with the reduction of SMN protein levels. Motor neuron degeneration and skeletal muscle atrophy are hallmarks of SMA, but it is unknown whether other mechanisms contribute to the spectrum of clinical phenotypes. Here, through a combination of physiological and morphological studies in mouse models and SMA patients, we identify dysfunction and loss of proprioceptive sensory synapses as key signatures of SMA pathology. We demonstrate that SMA patients exhibit impaired proprioception, and their proprioceptive sensory synapses are dysfunctional as measured by the neurophysiological test of the Hoffmann reflex (H-reflex). We further show that loss of excitatory afferent synapses and altered potassium channel expression in SMA motor neurons are conserved pathogenic events found in both severely affected patients and mouse models. Lastly, we report that improved motor function and fatigability in ambulatory SMA patients and mouse models treated with SMN-inducing drugs correlate with increased function of sensory-motor circuits that can be accurately captured by the H-reflex assay. Thus, sensory synaptic dysfunction is a clinically relevant event in SMA, and the H-reflex is a suitable assay to monitor disease progression and treatment efficacy of motor circuit pathology.

**One-sentence summary:** Sensory-motor circuit dysfunction involving impairment of proprioceptive synapses on motor neurons is a conserved pathogenic event and therapeutic target across animal models and humans with spinal muscular atrophy.

## INTRODUCTION

Studies in mouse models of human diseases are critical to gain insights into pathogenic mechanisms as well as to determine the efficacy and safety of potential treatments. However, determining whether knowledge acquired from studies in mice is applicable to patients can be challenging. This is especially the case in neurodegenerative disorders due to the difficulty of accessing neuronal circuits within the central nervous system in living patients. A prominent example of neurodegenerative diseases that affect motor circuits embedded in the spinal cord is spinal muscular atrophy (SMA). The dysfunction of sensory-motor circuits is a major pathogenic factor contributing to disease progression and severity in mouse models of SMA (Buettner et al., 2021; Fletcher et al., 2017; Mentis et al., 2011; Shorrock et al., 2019). However, whether the same mechanisms apply in SMA patients is currently unknown. This question is of particular clinical importance since the therapeutic landscape of SMA patients has changed dramatically with the introduction of three FDA-approved treatments (Mercuri et al., 2022). The current treatments increase life expectancy but do not fully restore motor function, calling for better understanding of disease mechanisms and development of additive therapies.

SMA is caused by homozygous inactivation of the survival motor neuron 1 gene (*SMN1*) and retention of at least one copy of the paralog gene *SMN2*, which is hypomorphic and results in ubiquitous deficiency of the SMN protein (Lefebvre et al., 1995). SMA is a childhood-onset disorder characterized by a varying degree of severity, which correlates with the copy number of *SMN2* genes (Mercuri *et al*., 2022). Historically, the severe form of SMA (Type 1) resulted in death by approximately two years after birth (Mercuri et al., 2012; Tizzano and Finkel, 2017). Individuals affected by the intermediate (Type 2) form of the disease never walk independently (Mercuri et al., 2018b). Type 3 SMA patients are more mildly affected, yet present with significant motor impairments even when ambulatory (Pera et al., 2017). The hallmarks of the disease are loss of spinal motor neurons, skeletal muscle atrophy, and severe motor impairment (Sumner and Crawford, 2018; Tisdale and Pellizzoni, 2015). Recently, three different SMN-inducing therapies have been approved for treatment of SMA patients (Erdos and Wild, 2022; Mendell et al., 2017; Mercuri et al., 2018a). Despite some clinical efficacy, especially in extending survival of Type 1 patients, significant motor impairments still persist in the majority of treated patients.

SMA affects primarily proximal and axial muscles in humans (Brogna et al., 2020; Deymeer et al., 1997), and studies in animal models of the disease revealed that functional disruption of sensory-motor circuits controlling these muscles contributes to motor deficits (Buettner et al., 2021; Fletcher et al., 2017; Imlach et al., 2012; Mentis et al., 2011; Shorrock et al., 2018; Simon et al., 2016). Accordingly, the dysfunction and loss of proprioceptive sensory synapses onto motor neurons is one of the most conserved, early pathogenic events across different mouse models of SMA (Buettner et al., 2021; Carlini et al., 2022; Cerveró et al., 2018). In the SMNΔ7 mouse model of SMA (Le et al., 2005), proprioceptive synapses release less glutamate, resulting in the reduction of Kv2.1 channel expression on motor neurons, effectively rendering them unable to fire repetitively (Fletcher et al., 2017). This ultimately contributes to impaired muscle contraction and diminished limb movements. Further underlying the significance of sensory synaptic defects in SMA, restoration of proprioceptive synaptic function improves motor behavior in both mouse and fly models of SMA (Fletcher et al., 2017; Imlach et al., 2012; Lotti et al., 2012; Simon et al., 2019). Importantly, however, it is unknown whether similar mechanisms involving proprioceptive synapses and consequent functional deficits in motor neuron output are at play in SMA patients and can be effectively targeted by current treatments.

Here, we utilized morphological and functional assays to determine whether proprioceptive sensory dysfunction characteristic of SMA mouse models is a disease feature in humans with SMA. We show that both SMNΔ7 mice and Type 3 ambulatory SMA patients display a marked reduction in the amplitude of the Hoffman reflex (H-reflex) compared to normal controls, demonstrating that proprioceptive sensory synapses are dysfunctional in SMA. This is further supported by evidence for the loss of proprioceptive synapses and dysregulated expression of Kv2.1 in SMA motor neurons from postmortem tissue of Type 1 SMA patients. Lastly, we show that significant improvement in the H-reflex correlates with an increase in walking distance and a concomitant reduction in fatigability in Type 3 SMA patients treated with an SMN-inducing therapy. Taken together, our results demonstrate that proprioceptive synaptic dysfunction is a mechanistically conserved pathogenic event in both mouse models and SMA patients, which has the potential to modify disease severity if successfully targeted therapeutically.

## RESULTS

### Ambulatory Type 3 SMA patients have defective proprioception

Despite strong evidence of proprioceptive sensory dysfunction in mouse models, it is unknown whether SMA patients exhibit any sensory deficits. Thus, we designed several sensitive assays to investigate whether proprioception is affected in ambulatory SMA patients. We first measured the angular difference in detection of passive movement of the knee at a constant speed of 0.5°/s, while the participants were both blind folded and aurally isolated (Fig. 1A). Three Type 3 SMA patients and three control participants were included in this study (see Methods). The test started from an initial position of the leg at 45° from the horizontal plane and then randomly selected flexion or extension movements were imposed to the knee joint (Fig. 1B). Remarkably, we found that all three SMA participants detected the movement at greater angles from the initial position compared to controls (Fig. 1C), indicative of impaired proprioception in SMA. Furthermore, we found that the deficit in detecting movement – illustrated by the higher detection angle – was proportional to the level of motor impairment for each participant as measured by the Hammersmith Functional Motor Scale Expanded (HFMSE) score (the lower the score, the greater the impairment; Fig. 1C). These results provided the first indication that proprioception is impaired and correlates with disease severity in Type 3 SMA patients.

**Figure 1.**
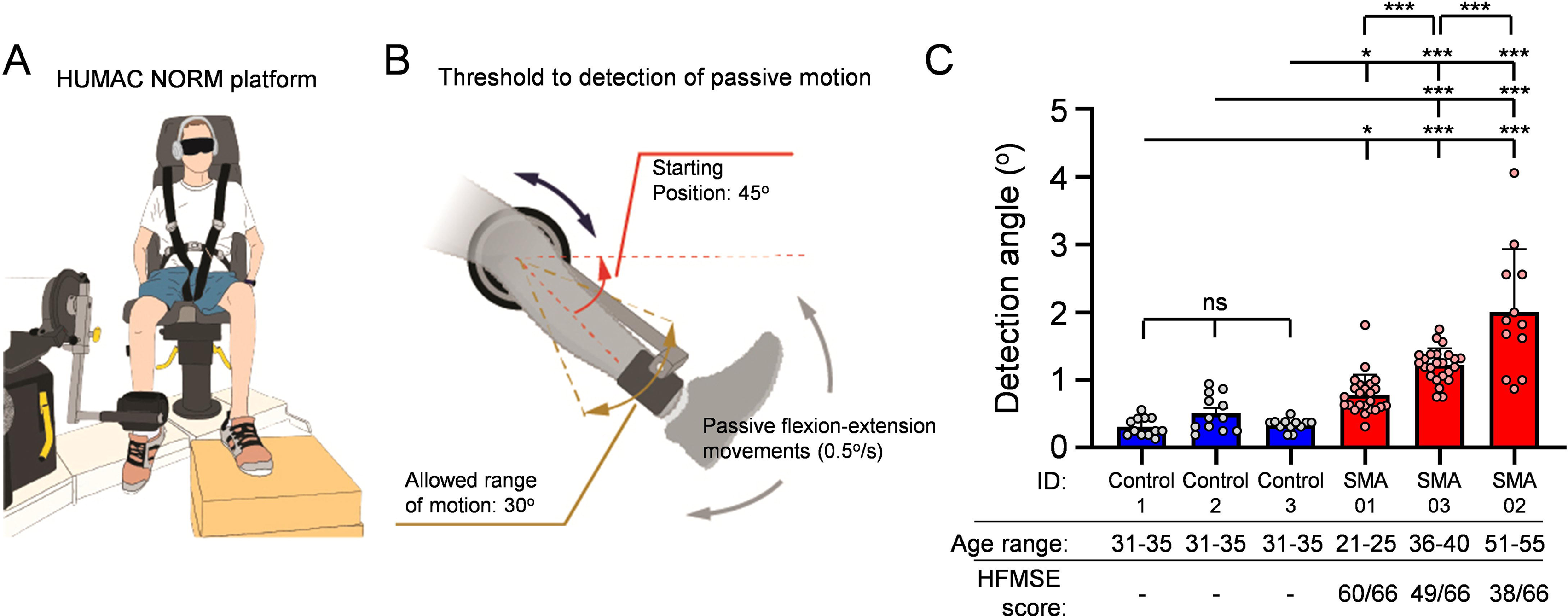
Proprioceptive dysfunction in SMA patients. (**A**) Drawing of the testing platform. The Humac Norm was used to assess proprioception in SMA patients and controls. This platform allows for precise passive movement of the knee joint at minimal speed of 0.5°/s. (**B**) Protocol details of the proprioceptive assessment test. The initial position of the leg was at 45° from the horizontal plane. Randomly selected flexion or extension movements were imposed in the knee joint at a speed of 0.5°/s and a maximum range of motion of 30°. (**C**) Deficits in proprioception vs Hammersmith Functional Motor Scale Expanded (HMFSE) motor score in three SMA patients (red bars) and three control participants (blue bars). Each dot per participant represent the result from an individual trial. The HFMSE scores for the SMA participants are also shown (max value 66). Statistical comparisons were performed between each SMA patient to each participant from the control group. Statistical significance was calculated using οrdinary one-way ANOVA with Tukey’s multiple *post hoc* comparisons test; *** p<0.001, * p<0.05; ns: no significance.

### The H-reflex is affected in Type 3 SMA patients

To measure directly the functionality of sensory-motor synapses, we performed the H-reflex. We employed three Type 3 SMA participants who never received any SMN-inducing therapy, three Type 3 SMA patients that were under nusinersen treatment (between 9-12 months at the time of our investigation), and seven control participants (Table 3). To quantify excitatory sensory drive to the spinal cord we measured the strength of the monosynaptic reflex circuit with the H-reflex test from the soleus muscle (Fig. 2A). In all groups, the H-reflex was observed at the lowest stimulation intensities (Suppl. Fig. 1A), while the M-response (short for “Muscle”; caused by direct stimulation of motor neuron axons) was evident at higher stimulation intensities with a concomitant reduction of the H-reflex until its abolition (Fig. 2B and Suppl. Fig. 1A). The amplitude of the M-response was reduced by ∼60% in participants with SMA compared to controls (Fig. 2B,C), likely reflecting neuromuscular junction (NMJ) denervation. Remarkably, the amplitude of the H-reflex was reduced by ∼90% in naïve SMA individuals (Fig. 2B,D). Importantly, these absolute differences were reflected in the normalized H/M amplitude ratio (Fig. 2E), eliminating any confounds due to possible differences in muscle mass. The latency for either M or H responses was not significantly different (Suppl. Fig. 1B,C). These findings demonstrate that the spinal monosynaptic reflex is affected in Type 3 SMA patients, indicating proprioceptive sensory dysfunction in humans. Interestingly, Type 3 SMA patients who were treated with nusinersen for ∼1 year showed greater H-reflex amplitudes and significantly improved H/M ratios (Fig. 2B,D,E), but no difference in the amplitudes of the M responses (Fig. 2C) compared to untreated patients. Moreover, nusinersen-treated SMA patients performed significantly better in the six-minute walking test (6MWT) (Fig. 2F) and displayed less fatigability (Fig. 2G) than naïve SMA patients. These results highlight a correlation between increased H-reflex amplitude and improved motor function in Type 3 SMA patients following nusinersen treatment, which may reflect increased sensory-motor circuit function and preservation of proprioceptive synapses onto surviving motor neurons.

**Figure 2.**
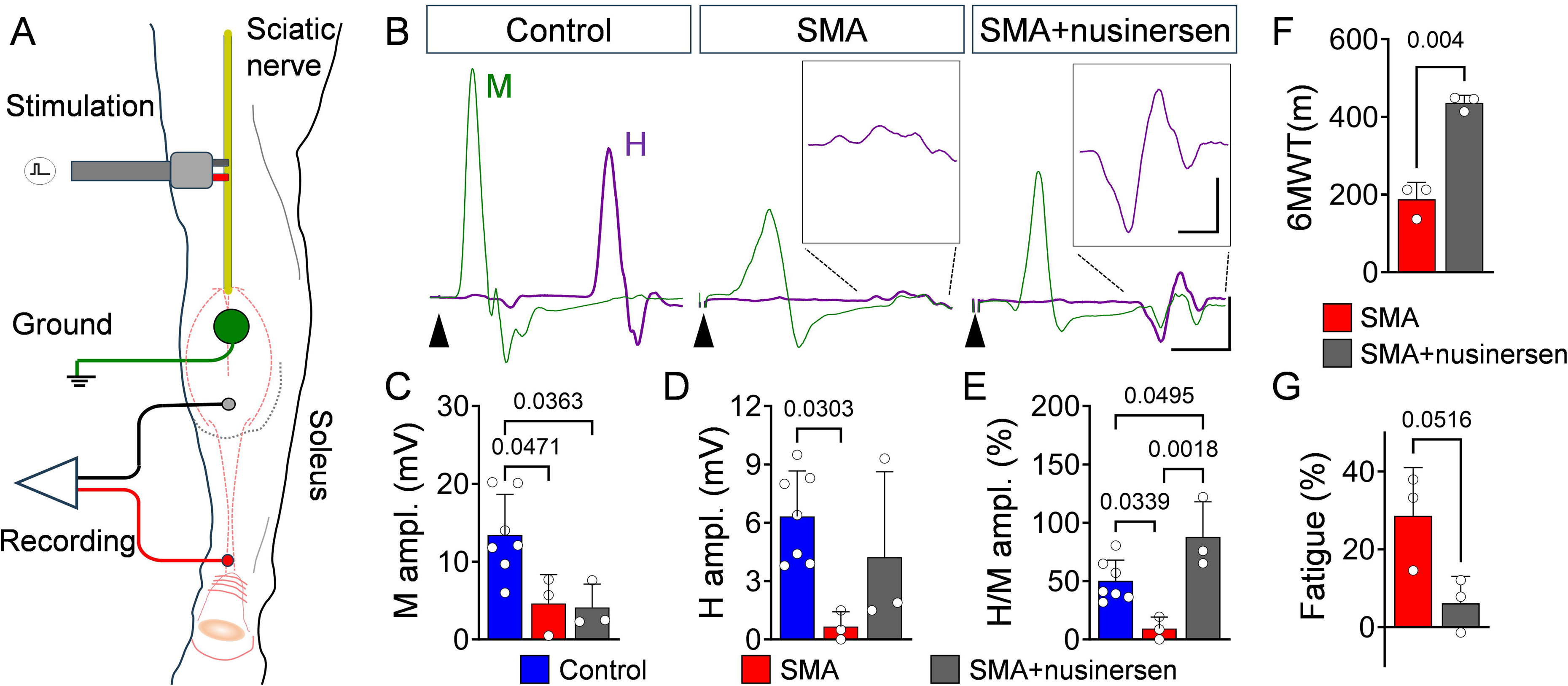
H-reflex is reduced in Type 3 SMA patients and improved following nusinersen treatment. (**A**) Schematic showing the location of the electrodes for the H-reflex recorded from soleus muscles in human participant. (**B**) EMG recordings from soleus muscle following stimulation of the tibial nerve in control, untreated and nusinersen-treated SMA patients. Superimposed traces for maximum M-(green) and H-(magenta) responses are shown. Arrowheads indicate stimulus artefacts. Scale bar: 2mV, 10ms. Insets show magnified H-reflex responses. Scale bar: 1mV, 5ms. M-response amplitude (**C**), H-reflex amplitude (**D**) and H/M amplitude ratio (**E**) for control (N = 7) and SMA-untreated (N = 3) and nusinersen-treated (N = 3) SMA Type 3 patients. (**F**) 6-minute-walk-test (6MWT) and (**G**) fatigability of untreated (N = 3) and nusinersen-treated (N = 3) SMA Type 3 patients. Data represent means and SD. Statistical analysis was performed using Welch’s test for (F), unpaired *t* test for (G), one-way ANOVA with Tukey multiple comparison test (C-E).

### The H-reflex is severely affected in sensory-motor circuits resistant to neurodegeneration

Proximal muscles are more vulnerable than distal ones in SMA patients (Brogna et al., 2020; Deymeer et al., 1997) as well as in SMNΔ7 mice (Ling et al., 2012; Mentis et al., 2011). Accordingly, upper lumbar motor neurons (L1-L2) exhibit greater muscle denervation than caudal lumbar (L4-L5) motor neurons, which do not degenerate in SMA mice (Fletcher et al., 2017; Kong et al., 2021; Mentis et al., 2011). Since mouse models that faithfully recapitulate milder SMA pathology are unavailable, we investigated neurodegeneration-resistant circuits in the caudal lumbar segments of SMNΔ7 mice as a proxy for motor circuits of Type 3 SMA patients. We employed the *ex vivo* spinal cord-hindlimb preparation and performed the H-reflex test from the tibialis anterior (TA) muscle in mice at P10 (Fig. 3A). Similar to humans, the H-reflex was evident at the lowest stimulation intensities of the common peroneal (CP) nerve (Suppl. Fig. 2A). At progressively greater stimulation intensities, the H-reflex was proportionally reduced while the M response increased in amplitude (Suppl. Fig. 2A). We confirmed the nature of the glutamatergic transmission of the H-reflex since pharmacological exposure to 10μM NBQX (AMPA receptor blocker) abolished the H-reflex leaving the cholinergic M-response relatively unaffected (Suppl. Fig. 2B). During early development, the H-reflex was also labile and prone to depression with eventual abolition following high frequency stimulation (10Hz), while the M-response was unaffected (Suppl. Fig. 2C). Importantly, SMA mice exhibited a marked reduction in both maximum M and H responses relative to normal controls (Fig. 3B-D), which was also reflected in reduction of the H/M ratio (Fig. 3E). Interestingly, the H-reflex amplitude was reduced by ∼85% (Fig. 3D), whereas the M-response was reduced by ∼45% in SMA (Fig. 3C). No differences were observed in the latency of either the M or H responses (Suppl. Fig. 2D), suggesting that the conduction velocities are not affected in either proprioceptive neurons or motor neurons. Akin to findings in milder Type 3 SMA patients, these results demonstrate that the H-reflex is severely affected in sensory-motor circuits of SMA mice that are resistant to neurodegeneration.

**Figure 3.**
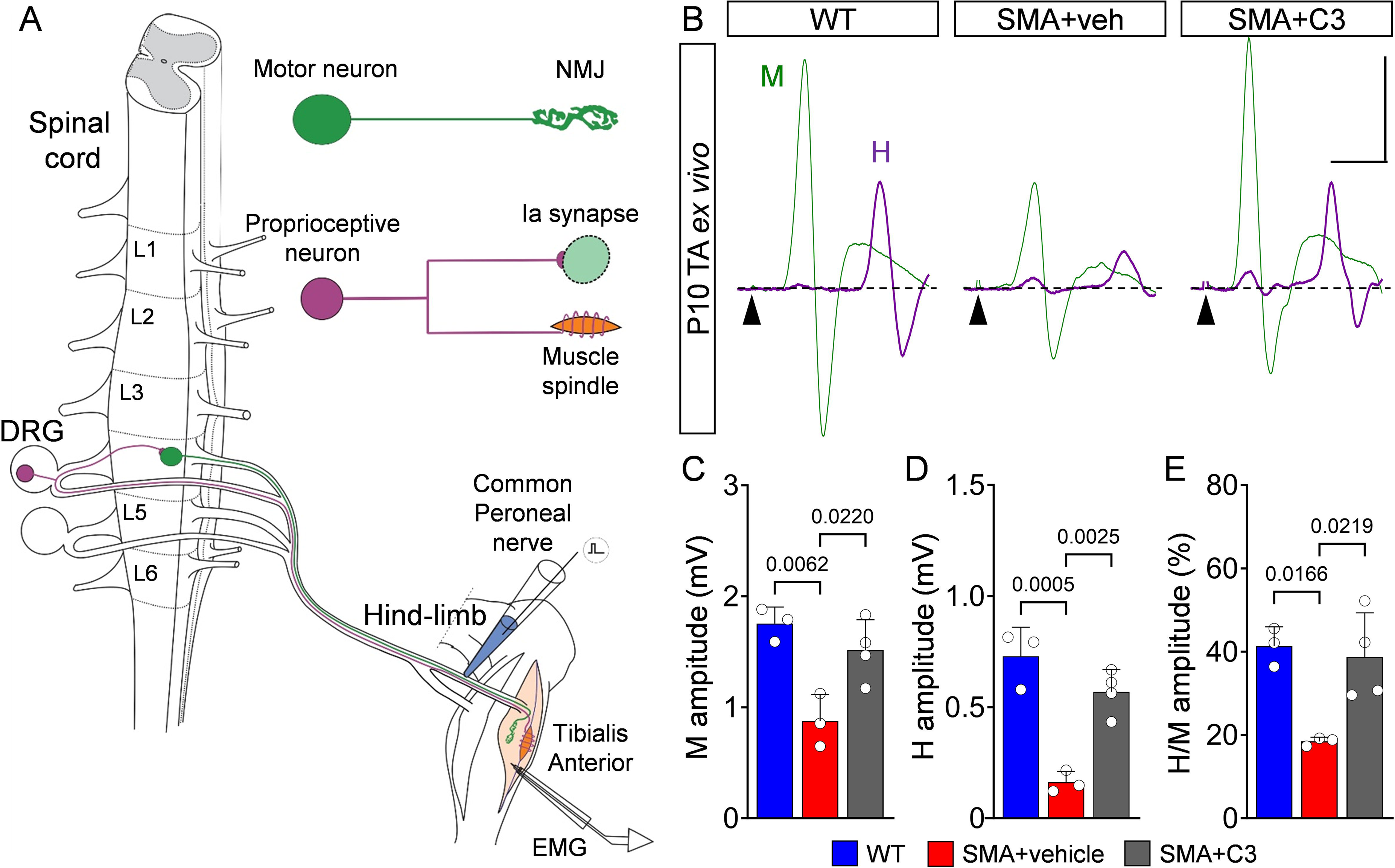
H-reflex is reduced in neonatal SMA mice. (**A**) Experimental setup to measure H-reflex using the spinal cord-hindlimb *ex vivo* preparation. The stretch reflex circuit consists of a motor neuron (green) and its neuromuscular junction (NMJ) synapse with its muscle, as well as a proprioceptive neuron (magenta) originating from a muscle spindle and making contact with a motor neuron via an Ia synapse. The common peroneal (CP) nerve was stimulated with an *en passant* electrode, while EMG was recorded from the tibialis anterior (TA) muscle. (**B**) TA EMG recordings following stimulation of the CP nerve in WT, vehicle-treated SMA and C3-treated SMA mice at P10. Superimposed traces for maximum M-(green) and H-(magenta) responses are shown. Arrowheads indicate stimulus artefacts. Scale bar: 0.5mV, 5ms. M-response amplitude (**C**), H-reflex amplitude (**D**) and H/M amplitude ratio (**E**) from WT (N = 3 mice) and SMA+vehicle (N = 3) and SMA+C3 (N = 4) mice at P10. Data represent means and SD. Statistical analysis was performed using an one-way ANOVA with Tukey multiple comparison test (C-E).

### Pharmacological upregulation of SMN improves the H-reflex and sensory-motor circuit function in SMA mice

We sought to investigate whether upregulation of SMN would improve the H-reflex and sensory-motor circuit function in SMA mice. To do so, we treated SMNΔ7 mice with the SMN-inducing compound C3 (Naryshkin et al., 2014), an analogue of risdiplam which was previously shown to extend lifespan and significantly improve the overall SMA phenotype in this mouse model (Naryshkin et al., 2014). Accordingly, SMNΔ7 mice treated daily from P0 with 3mg/kg of C3 displayed strong progressive improvement in righting (Fig. 4A), posture time (Fig. 4B), and weight gain (Fig. 4C). Importantly, we found that both M and H responses recorded from the TA muscle of C3-treated SMA mice at P10 were fully rescued (Fig. 3B-E). To investigate whether the beneficial effects of C3 treatment in sensory-motor circuits of SMA mice were maintained at later times, we examined the H-reflex in the flexor digitorum brevis (FDB) muscle following stimulation of the sciatic nerve *in vivo* and under general anesthesia at P30 (Fig. 4D). Motor neurons innervating FDB muscle are located in the L4-L6 spinal segments (Bácskai et al., 2014), and this muscle was chosen for its reliability in obtaining H-reflexes *in vivo* and vulnerability in SMA mice (Ling *et al*., 2012). We found that SMA-treated mice exhibited nearly normal M and H responses as well as H/M ratios compared to age-matched WT controls (Fig. 4E-H), while there were no differences in their corresponding latencies (Suppl. Fig. 3A,B). Together, these findings reveal that the behavioral benefit of treatment with an SMN-inducing drug correlates with improved H-reflex and sensory-motor circuit function in SMA mice.

**Figure 4.**
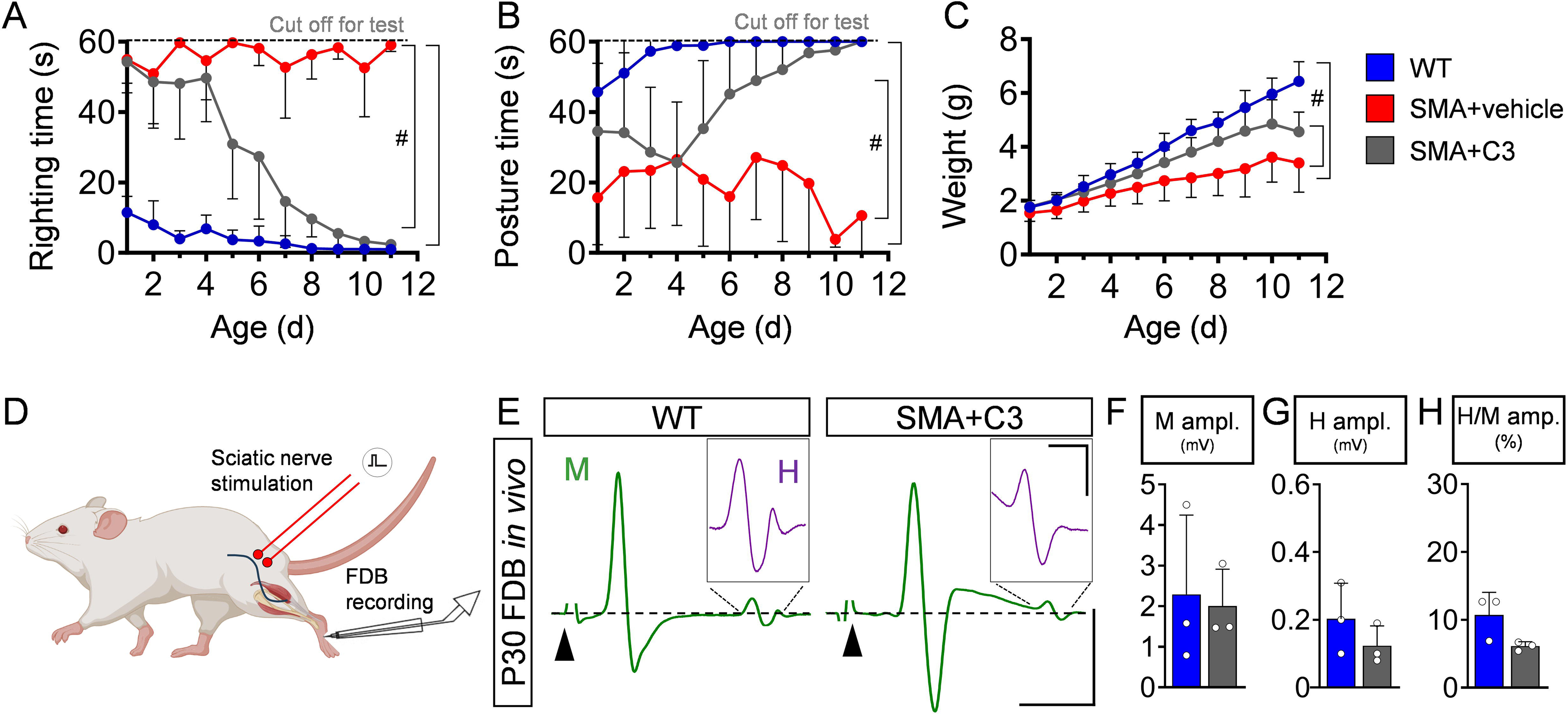
SMN therapy restores H-reflex in juvenile SMA mice. Righting time (**A**), posture time (**B**) and weight (**C**) of WT (blue, N = 12 mice), SMA+vehicle (red, N = 9), and SMA+C3 (grey, N = 14) mice. (**D**) Drawing of a mouse highlighting the approximate location for stimulation of the sciatic nerve (red electrode) and the approximate location of the EMG electrode for the flexor digitorum brevis (FDB) muscle at the sole of the hindpaw. (**E**) *In vivo* EMG recordings form FDB muscle following stimulation of the sciatic nerve in WT and SMA mice treated daily with C3 compound at P30. Traces for M- and H-responses are shown in green. Arrowheads indicate stimulus artefact. Scale bar: 1mV, 2ms. Insets show magnified H-reflex in magenta. Scale bar: 50μV, 0.5ms. M-response amplitude (**F**), H-reflex amplitude (**G**), and H/M amplitude ratio (**H**) from WT (N = 3) and SMA+C3 (N = 3) mice at P30. Data represent means and SD. Statistical analysis was performed using t-test (F,G,H) and two-way ANOVA with Tukey multiple comparison test (A,B,C). # indicates *P*˂0.05.

### Spinal sensory-motor circuits controlling hindlimb muscles become dysfunctional at late disease stages in SMA mice

The reduction in the H-reflex from L4-L5 sensory-motor circuits comprising degeneration-resistant motor neurons such as those controlling the TA muscles in SMNΔ7 mice prompted us to investigate their functional status in greater detail. To do so, we employed a modified version of the *ex vivo* spinal cord preparation (Özyurt et al., 2022) and recorded L4/L5 motor neuron responses from P10 WT and SMNΔ7 mice intracellularly, following suprathreshold proprioceptive sensory fiber stimulation (see Methods and Fig. 5A,B). We found reduced amplitude of excitatory postsynaptic potentials (EPSPs) in SMA motor neurons compared to their WT counterparts (Fig. 5C,D), with no change in their latency (Fig. 5E). Monosynaptically-mediated EPSPs were not correlated with changes in motor neuron input resistance (Fig. 5F). Similarly, there was no correlation between input resistance and the cross-section soma area for all recorded motor neurons (Fig. 5G). We also examined whether the reduction in the EPSP amplitude might reflect a reduction of the number of synapses impinging on motor neurons. However, we found no differences in the number of proprioceptive (VGluT1^+^) synapses on the soma of WT and SMA motor neurons (Fig. 5H,I) and only a minimal decrease on proximal dendrites (Suppl. Fig. 4A).

**Figure 5.**
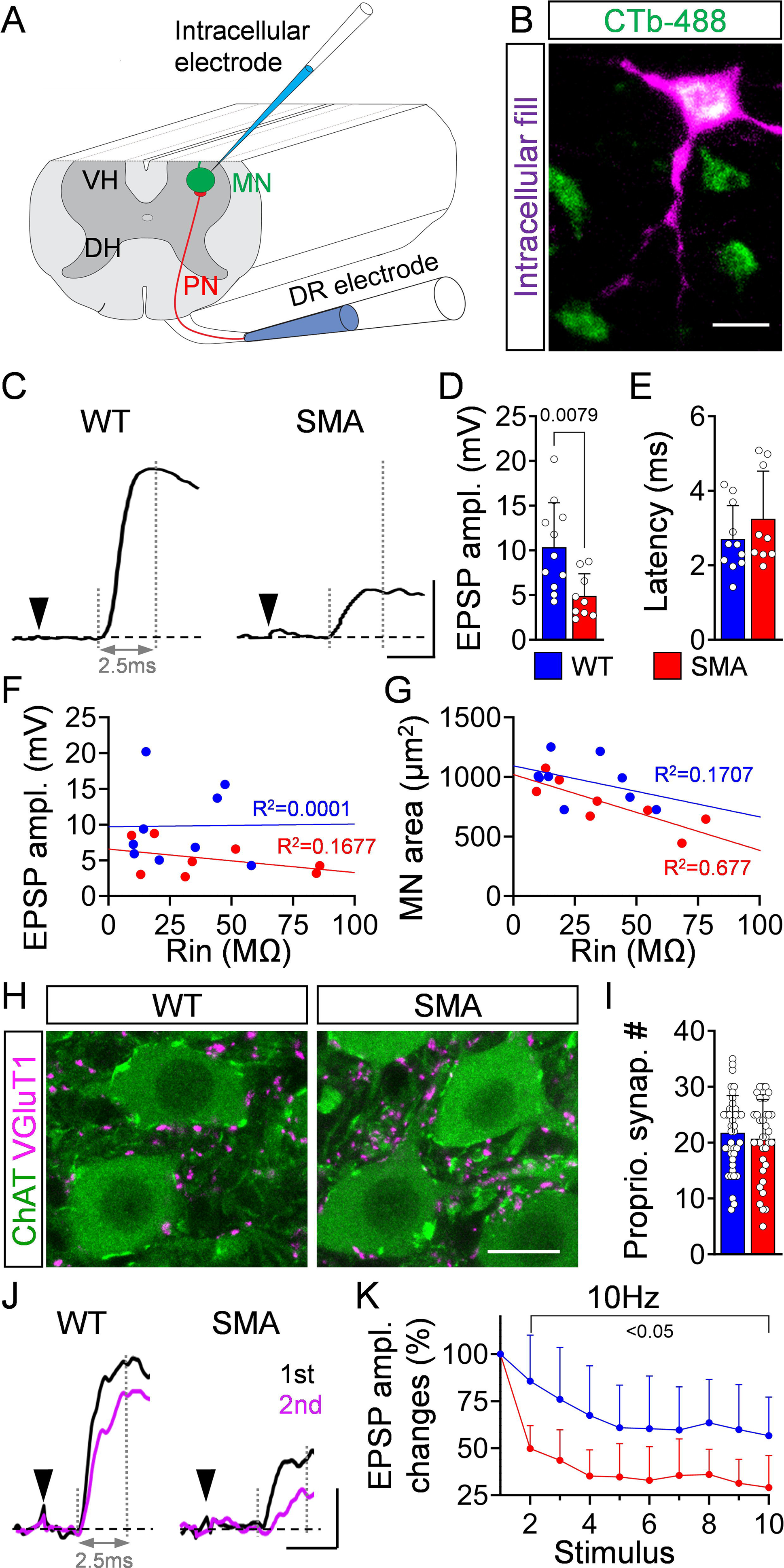
Proprioceptive neurotransmission is impaired in motor neurons innervating distal muscles at late stages of disease in SMA mice. (**A**) Schematic of the modified *ex vivo* spinal cord preparation in which the ventral funiculus was removed to aid visual access for patch clamp recordings. (**B**) Confocal image of CTb-488 (in green) motor neurons in a spinal cord preparation (L4/5 spinal segment). A motor neuron is shown in magenta which was intracellularly recorded, filled with Neurobiotin and visualized *post hoc*. Scale bar: 20µm. (**C**) Excitatory postsynaptic potentials (EPSPs) following supramaximal L5 dorsal root stimulation in WT and SMA mice at P10. Dotted vertical lines indicate the onset of the response and where the EPSP amplitude is measured (2.5ms after the onset). Scale bar: 5mV, 2ms. (**D**) peak amplitude and (**E**) latency of EPSPs in WT (*n* = 11 motor neurons) and SMA (*n* = 9) motor neurons at P10. At least N=4 mice used per genotype. Relationship between input resistance and peak EPSP amplitude (**F**) as well as input resistance and motor neuron soma area (**G**) from WT (*n* = 9) and SMA (*n* = 8) motor neurons at P10. (**H**) Confocal images of L5 motor neurons (ChAT, green) and proprioceptive synapses (VGluT1, magenta) from WT and SMA mice at P10. Scale bar: 20µm. (**I**) Number of VGluT1+ proprioceptive synapses onto the soma of L5 motor neurons from WT (*n* = 40 MNs, N = 4 mice) and SMA (*n* = 30 MNs, N = 3 mice) mice at P10. (**J**) First (black) and second (magenta) EPSP responses elicited in motor neurons after 10Hz dorsal root stimulation in P10 WT and SMA mice. Arrowheads indicate stimulus artefact. Vertical dotted line marks peak EPSP amplitude measured at 2.5ms after the onset of response. Scale bar: 2mV, 2.5ms. (**K**) Percentage changes in EPSP amplitude following 10 stimuli at 10Hz normalized to the first response for WT (*n* = 10) and SMA (*n* = 9) motor neurons at P10. Data represent means and SD. Statistical analysis was performed using Mann-Whitney test for (C, I), unpaired *t* test for (B, G), and simple linear regression (D, E).

To test the possibility that glutamate release may be the cause for the reduction in synaptic transmission, we performed dorsal root stimulation at different frequencies and measure the EPSP amplitudes between subsequent trials. We found no significant difference between WT and SMA mice at 0.1Hz (Suppl. Fig. 4C), and a ∼25% reduction at 1Hz in SMA motor neurons after several stimuli (Suppl. Fig. 4B,D). In contrast, at the more physiologically-relevant stimulation frequency of 10Hz, there was a strong reduction (∼50%) of the EPSP amplitude in SMA mice compared to a smaller reduction (∼13%) in WT mice as early as the 2^nd^ stimulus (Fig. 5J,K). These results suggest that the ∼50% reduction in synaptic transmission occurs via presynaptic mechanisms through impairment of glutamate release by proprioceptive synapses onto degeneration-resistant motor neurons at later stages of the disease.

We have previously reported that dysfunction of proprioceptive synapses has profound effects on the output of vulnerable motor neurons in SMA mice (Fletcher et al., 2017). Therefore, we tested whether this applies to degeneration-resistant SMA motor neurons innervating the TA muscle through analysis of their intracellular properties at P10 in SMNΔ7 mice. We found no significant differences in resting membrane potential (Suppl. Fig. 5A). In contrast, the passive membrane properties of input resistance (R_in_) and time constant (τ) revealed a significant increase in SMA neurons compared to controls (Fig. 6A-C). This increase was unlikely due to changes in soma size because the capacitance did not differ (Suppl. Fig. 5B). Furthermore, the rheobase was inversely and proportionally reduced in SMA motor neurons (Fig. 6A,D), pointing to changes in the distribution of ion channels at the cell membrane (specific membrane resistivity). Lastly, we examined the ability of TA motor neurons to fire repetitively by injecting steps of current and found that SMA motor neurons exhibited significantly lower firing frequencies (Fig. 6E,F).

**Figure 6.**
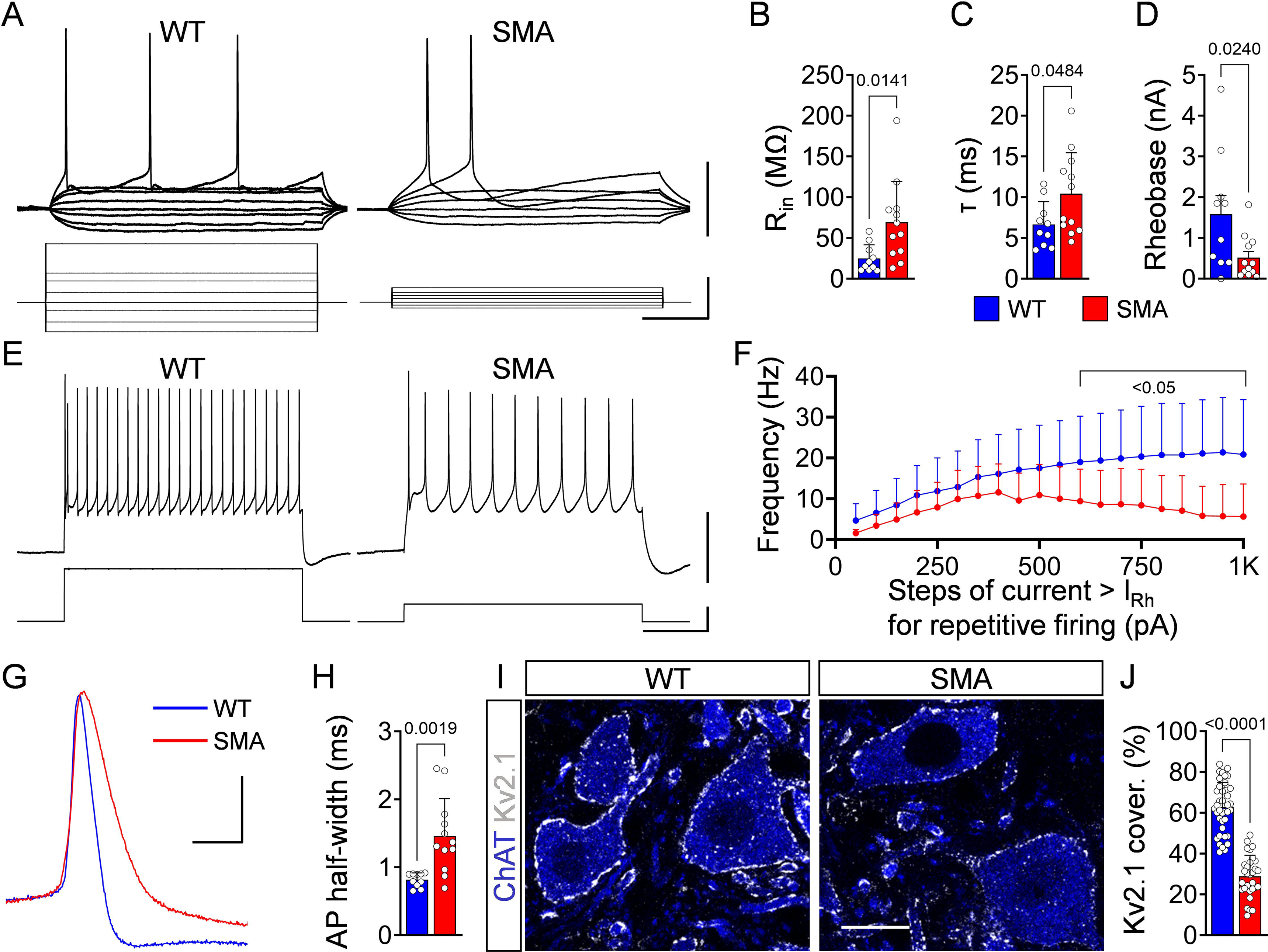
Motor neurons innervating distal muscles exhibit signs of dysfunction in symptomatic SMA mice. (**A**) Membrane responses to current injections in a WT and SMA L5 motor neurons at P10. Scale bars: 20mV, 400pA, 100ms. (**B**) Input resistance, (**C**) Time constant and (**D**) Rheobase for WT (*n* = 10 motor neurons) and SMA (*n* = 12) motor neurons at P10. At least N=4 mice used per genotype. (**E**) Intracellular responses (top) showing repetitive firing at the maximum frequency attained during current injection (bottom) and (**F**) Frequency-to-current relationship for WT (*n* = 9) and SMA (*n* = 12) at P10. Scale bar: 40mV, 500pA, 200ms. (**G**) Superimposed action potentials during steady-state firing following current injection in a WT (blue) and SMA (red) motor neuron at P10. Scale bar: 20mV, 2ms. (**H**) Duration at half-width of action potentials for WT (*n* = 10) and SMA (*n* = 12) motor neurons. (**I**) Single-optical-plane confocal images of L5 motor neurons (ChAT, blue) expressing Kv2.1 channels (white) from P10 WT and SMA mice. Scale bar: 20µm. (**J**) Percentage somatic coverage of Kv2.1 expression in WT (n = 40 MNs, N = 4 mice) and SMA motor neurons (n = 30 MNs, N = 3 mice). Data represent means and SD. Statistical analysis was performed using unpaired *t* test for (B,C,J), Mann-Whitney test for (D, F) and Welch’s *t* test for (H).

A key molecular mechanism for the reduced firing ability of vulnerable SMA motor neurons is the reduction of Kv2.1 channel expression, a major contributor of the delayed rectification of motor neuron’s action potentials (AP) (Fletcher et al., 2017). Therefore, we investigated several characteristics of the APs in WT and SMA motor neurons innervating the TA. We found no significant difference in the AP amplitude (Suppl. Fig. 5C), but a marked reduction in voltage threshold (V_thr_) in SMA motor neurons compared to controls (Suppl. Fig. 5D). Importantly, the half-width of AP was significantly slower in the repolarizing phase in SMA motor neurons (Fig. 6G,H) and immunoreactivity against Kv2.1 channels revealed a significant >50% reduction in their coverage of soma membrane (Fig. 6I,J). These results indicate that even SMA motor neurons that are resistant to neuronal death become progressively dysfunctional during the course of the disease.

### Proprioceptive synapses and Kv2.1 channels are reduced in motor neurons from SMA patients

While we observed dysfunction of proprioception and monosynaptic reflexes in humans with SMA, it is not known whether related pathogenic features such as loss of proprioceptive VGluT1^+^ synapses and reduced expression of Kv2.1 channels in spinal motor neurons that are found in SMA mouse models also occur in humans with SMA. To address this, we investigated morphological signatures of sensory-motor circuit pathology in post-mortem spinal cords from Type 1 SMA patients and aged-matched controls.

First, we quantified the number of motor neurons, the loss of which is a hallmark of severe SMA in both humans and mice. Remarkably, we found that both α and γ motor neurons were significantly reduced in SMA spinal cords compared to controls (Fig. 7A-C). Despite limitations in tissue collection did not allow us to determine the identity of the specific spinal segments, the size of the surviving motor neurons in SMA patients was significantly smaller compared to controls (Fig. 7D). The finding that ∼50% of the motor neurons survive until disease end stage and could be engaged in alleviating the severity of symptoms has important implications for SMA therapy.

**Figure 7.**
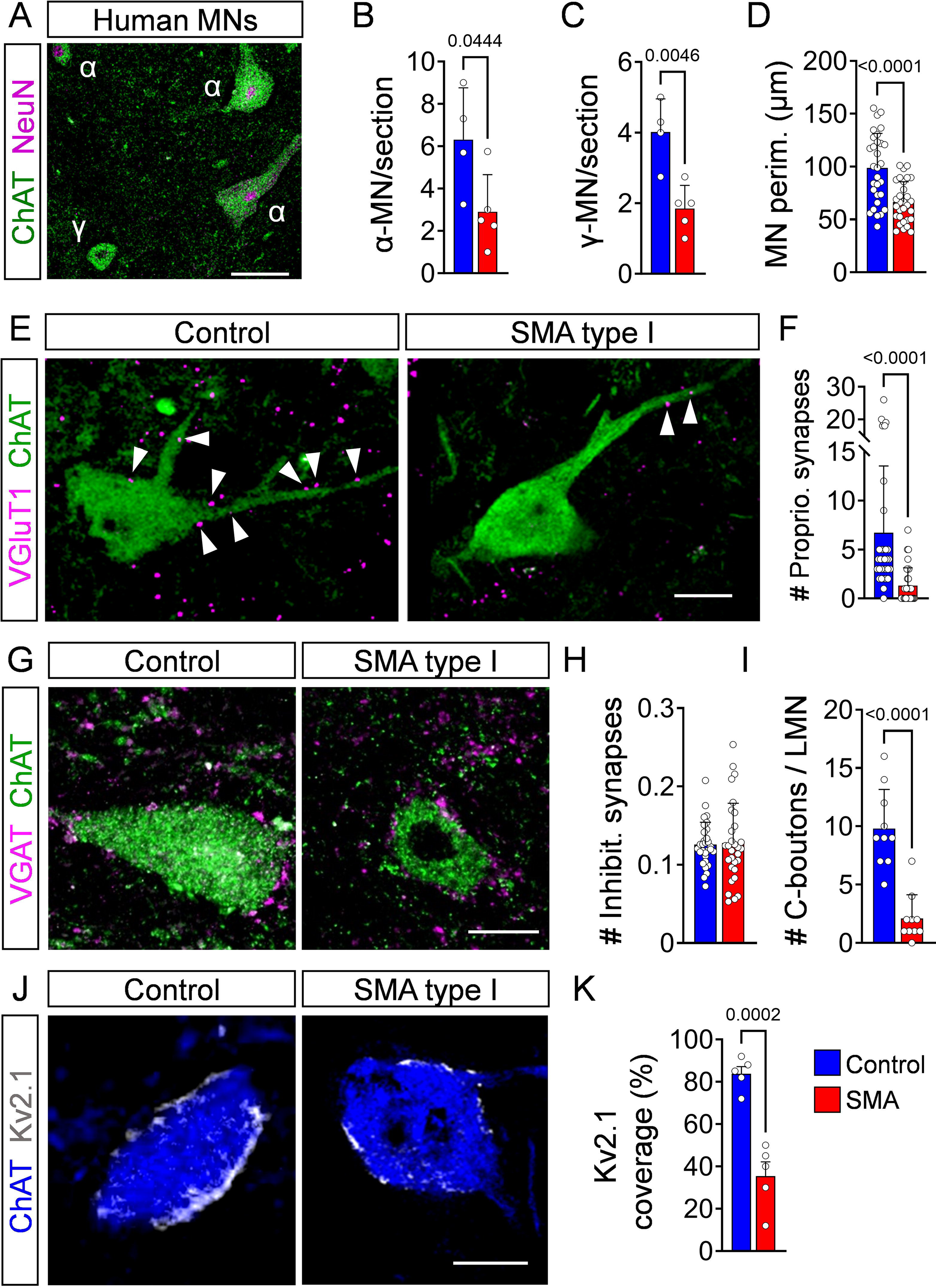
Proprioceptive synapses and Kv2.1 channels are reduced in motor neurons from Type I SMA patients. (**A**) Confocal image of α-motor neurons (MNs) (ChAT+, NeuN+) and γ-motor neurons (ChAT+, NeuN-) in a human control spinal cord. Scale bar: 50µm. Number of thoracic α-motor neurons (**B**) and γ-motor neurons (**C**) per spinal cord section in control (N = 4 subjects) and Type I SMA patients (N = 5 patients). (**D**) Perimeter of thoracic motor neuron soma in control (*n* = 30 MNs, N = 3 subjects) and Type I SMA spinal cords (*n* = 30 MNs, N = 3 patients). (**E**) Confocal images of motor neurons (ChAT; green) with proprioceptive synapses (VGluT1; magenta) from postmortem human spinal cord sections from a control and Type I SMA patient. Arrowheads point to proprioceptive synapses. Scale bar: 20µm. (**F**) Number of proprioceptive synapses on spinal motor neurons from control (*n* = 34 MNs, N = 3 subjects) and SMA type I patients (*n* = 32 MNs, N = 4 patients). (**G**) Inhibitory synapses (VGAT; magenta) on motor neurons (ChAT; green) from a control and SMA Type I patient. Scale bar: 20µm. (**H**) Number of inhibitory synapses on motor neurons from control (*n* = 30 MNs, N = 3 subjects) and SMA Type I patients (*n* = 30 MNs, N = 3 patients). (**I**) Number of C-boutons onto lumbar lateral motor neurons of control (*n* = 10 MNs, N = 1 subject) and Type I SMA patient (*n* =10 MNs, N = 1 patient). (**J**) Single-optical-plane confocal images of motor neurons (ChAT; blue) expressing Kv2.1 channels (white) from a control and SMA Type I patient. Scale bar: 20µm. (**K**) Percentage somatic coverage of Kv2.1 expression in control (*n* = 5 MNs, N = 2 subjects) and SMA Type I patients (*n* = 5 MNs, N = 2 patients) lumbar motor neurons. Data represent means and SD. Statistical analysis was performed using Welch’s *t* test for (B, C, F, I, K) and unpaired *t* test for (D, H).

Next, we quantified the number of VGluT1^+^ synapses on the soma of motor neurons. VGluT1 also marks corticospinal synapses (Lodato et al., 2014; Persson et al., 2006) on human and non-human primate motor neurons (Baldissera and Cavallari, 1993; de Noordhout et al., 1999; Jo et al., 2023; Lemon, 2008; Palmer and Ashby, 1992), but not rodent motor neurons (Alstermark et al., 2004; Gu et al., 2017; Yang and Lemon, 2003). However, these corticomotoneuronal-VGluT1^+^ synapses develop late, at ∼8-10 months after birth in non-human primates (Lawrence and Hopkins, 1976) and a year after birth in humans (Lundberg, 1979; Matijević et al., 2013). Thus, all VGluT1^+^ synapses are considered of proprioceptive origin during early human development. Remarkably, we found a significant reduction in VGluT1^+^ synapses on motor neurons of Type 1 SMA patients compared to age-matched controls (Fig. 7E,F). The postmortem interval (PMI) did not play a significant role in this analysis (Suppl. Fig. 6A) and VGluT1 synaptic coverage was significantly reduced on both medially-located lumbar motor neurons (MMC) - which innervate axial muscles – and laterally-located motor neurons (LMC) - which innervate distal muscles (Suppl. Fig. 6B,C). Interestingly, we found that the number of cholinergic C-boutons on lumbar SMA motor neurons was also reduced (Fig. 7G,I), while the number of inhibitory synapses (marked by the vesicular transporter, VGAT) did not change between control subjects and SMA Type 1 patients (Fig. 7G,H). Lastly, we quantified the percentage coverage of Kv2.1 on the soma of motor neurons and found a significant ∼60% reduction in SMA motor neurons (Fig. 7J,K).

Collectively, these findings indicate that key cellular and synaptic features of sensory-motor circuit pathology are conserved in severe mouse models and Type 1 SMA patients.

## DISCUSSION

This study reveals that impaired proprioceptive-mediated neurotransmission is a conserved pathogenic event that contributes to disease progression in both mouse models and patients affected by SMA. We show that the H-reflex is a reliable neurological test to assess the function of sensory-motor circuits in both animal models and ambulatory SMA patients. Moreover, through morphological analysis of postmortem spinal cords from severe Type 1 SMA patients, we show that the ∼50% of motor neurons that survive until disease end stage exhibit widespread loss of sensory synapses and a concomitant reduction in Kv2.1 channel expression. Importantly, the same defects occur in severe SMA mice and are responsible for reduced motor neuron firing that contributes to impaired muscle contraction. Lastly, we document that improved motor function upon treatment with SMN-inducing drugs such as nusinersen in SMA patients or the risdiplam-analogue C3 in SMA mice correlates with a strong increase in the H-reflex amplitude. Thus, the H-reflex could be used as a reliable functional assay for monitoring both progression and therapeutic targeting of sensory-motor circuit pathology in SMA.

Our study provides a first-in-human demonstration that SMA patients exhibit compromised proprioception compared to healthy individuals. Our first indication that proprioception is impaired in humans with SMA comes from the conscious joint position sense experiments. This test is different from the H-reflex test in that conscious impairment in proprioception includes ascending pathway through dorsal column and various brain structures, from the cuneate and gracilis nuclei to the sensory cortex (Dykes et al., 1982; Nyberg and Blomqvist, 1982; Salles et al., 2011). It is currently unclear whether these brain pathways are affected in SMA patients and contribute to affect proprioception together with the dysfunction of sensory-motor synapses which we have uncovered with the H-reflex. Accordingly, we found that ambulatory SMA patients exhibited marked reductions in the H-reflex amplitude, which was more pronounced than that of the M-response. This finding indicates that central sensory synapses are more affected than NMJs, which is supported by the analysis of the H/M amplitude ratio. Our results may also help better explain the results of an earlier electromyographic study in which the absence of H-reflexes in patients suffering from Werdnig-Hoffmann disease was attributed to the severe muscle denervation (Renault et al., 1983). Together with our recent report of two cases of SMA with sensory involvement (Oskoui et al., 2020), these results are well aligned with previous studies documenting dysfunction of proprioceptive synapses as a conserved disease feature in animal models (Buettner et al., 2021; Fletcher et al., 2017; Imlach et al., 2012; Mentis et al., 2011).

What are the mechanisms of proprioceptive dysfunction in SMA patients? Key insights into this question initially came from studies in SMNΔ7 mice, which faithfully model severe forms of SMA. At the onset of disease, proprioceptive synapses release less glutamate at the physiological frequencies (Fletcher et al., 2017). These synapses are then gradually lost from their corresponding motor neurons, resulting in reduced expression of the potassium channel Kv2.1, as we report here in human SMA motor neurons. In turn, the reduction of Kv2.1 markedly affects the ability of motor neurons to fire repetitively (Fletcher et al., 2017), and ultimately impairs muscle contraction. However, it is unknown whether this sequence of events applies to ambulatory Type 3 SMA patients. In the absence of reliable mouse models for milder forms of SMA, we sought to study degeneration-resistant motor neurons in the SMNΔ7 mice as a proxy for sensory-motor circuits that are less severely affected by SMN deficiency. We report here that, except for neuronal death, these motor neurons exhibit functional deficits akin to those found in vulnerable motor neurons but at later stages of disease. Thus, distinct SMA motor neurons become dysfunctional at different times during disease progression. These observations may help explain the slower, yet progressive motor decline in milder Type 3 SMA patients (Salort-Campana and Quijano-Roy, 2020).

Our analysis of postmortem spinal cord preparations from Type 1 SMA patients provides direct evidence that proprioceptive synapses are lost from human motor neurons similar to mouse models (Buettner et al., 2021; Fletcher et al., 2017; Shorrock et al., 2018; Simon et al., 2019). In further agreement with mouse studies (Buettner et al., 2021; Cerveró et al., 2018), the number of cholinergic synapses on the soma of motor neurons are also reduced in Type 1 SMA patients relative to controls. Proprioceptive and cholinergic synapses are key regulators of Kv2.1 channel’s expression in motor neurons (Deardorff et al., 2021). Strikingly, our study uncovered marked reduction of Kv2.1 channel expression in motor neurons from Type 1 SMA patients. Taken together, these findings provide strong evidence that the mechanisms of sensory-motor circuit pathology are conserved between mouse models and SMA patients.

What is the clinical significance of impaired proprioception in SMA? The test performed with the angular difference in detection of passive movement of the knee indicated that the proprioceptive impairment correlated with the HFMSE score and the length of disease in the three ambulatory Type 3 SMA participants. This implies that dysfunction in proprioception is progressive and can be quantified using the H-reflex. Furthermore, increased amplitudes of the H-reflex correlated with improved walking distance and reduced fatigability in all three SMA participants treated with nusinersen. Importantly, these observations parallel those from SMA mouse experiments in which improved motor function following treatment with an analogue of risdiplam was associated with an increase in H-reflex amplitude. While these are correlative observations, a causal link between proprioceptive dysfunction and disease pathogenesis was previously established in mouse model (Fletcher et al., 2017; Simon et al., 2019). Moreover, a recent study provided proof of principle that ambulatory SMA patients regain previously lost muscle strength and increase endurance following epidural spinal cord stimulation of proprioceptive afferents (Prat-Ortega, 2024).

Current therapies for SMA patients include an antisense oligonucleotide (nusinersen) (Wood et al., 2017), a splice-modifier molecule (risdiplam) (Hua et al., 2011; Naryshkin et al., 2014; Palacino et al., 2015) and SMN replacement by gene therapy (onasemnogene abeparvovec) (Kotulska et al., 2021; Passini et al., 2010), all of which aim to increase SMN protein via different mechanisms. Although all three therapies have demonstrated clinical efficacy (Aslesh and Yokota, 2022; De Vivo et al., 2019; Finkel et al., 2021; Finkel et al., 2018; Mendell et al., 2021; Mercuri et al., 2018b), it is widely accepted that none of these represent a cure (Mercuri et al., 2020; Reilly et al., 2022; Strauss et al., 2022a; Strauss et al., 2022b; Wirth, 2021) leaving many treated patients with significant motor deficits (Day et al., 2022; Mercuri et al., 2022; Nicolau et al., 2021). The response to current SMA therapies is variable and there is an increasing need for reliable measures of functional improvement to assess efficacy of treatment (Oskoui et al., 2023; Reilly et al., 2022).

Here, we show that Type 3 SMA patients exhibited significant improvements in the distance walked and fatigability following ∼12 months on nusinersen which were correlated with an increase in the H-reflex and H/M ratio, indicating that this test could capture functional improvement. Thus, we propose that analysis of the H-reflex should be implemented in clinical settings as a sensitive and reliable measure of sensory-motor circuit pathology in SMA patients. The H-reflex could monitor disease progression, assess efficacy of treatment, and identify potential deleterious effects of therapies (Van Alstyne et al., 2021). H-reflex tests can be performed in different muscles of both upper and lower limbs in humans (Zehr, 2002) as in patients with diabetic disorders (Kababie-Ameo et al., 2023). Upper limb muscles can be used for Type 2 SMA patients who are wheelchair bound, and lower limb muscles in ambulatory Type 3 SMA patients, as in our study.

In summary, we conclude that proprioceptive synaptic dysfunction is a conserved pathogenic event in humans and mouse models and propose that proprioceptive sensory neurotransmission should be targeted therapeutically in SMA.

## MATERIALS AND METHODS

### Study design

Sample size and rules for stopping data collection was determined by previous experience and preliminary data. All data was included if the experiment was technically sound (perfusion, tissue preparation, staining, recordings, imaging etc.). The endpoints for animals were selected by previous experiments and literature references. Each experiment was at least three times replicated with a few exceptions of human tissue due to availability limitations, as described below. The research objects were to investigate proprioceptive dysfunction in SMA patients and autopsy tissue as well as in a severe SMA mouse model. The mice were randomized to treatment group, and the investigators who assessed the behavioral, histological and electrophysiological outcomes were blinded to the treatment group. The identification of the SMA patients and that of control participants used here, are arbitrary and do not reflect a patient identification number.

### Animal procedure

Breeding and experiments were performed in the animal facilities of Columbia University, New York, USA and Leipzig University, Germany according to National Institutes of Health Guidelines on the Care and Use of Animals and approved by the Columbia Animal Care and Use Committee (IACUC) for Columbia University or European (Council Directive 86/609/EEC) and German (Tierschutzgesetz) guidelines for the welfare of experimental animals and the regional directorate (Landesdirektion) of Leipzig for Leipzig University.

Mice were housed in a 12h/12h light/dark cycle with access to food and water *ad libitum*. The original breeding pairs for SMNΔ7 (*Smn*^+/-^; *SMN2*^+/+^; *SMNΔ7*^+/+^) mice on FVB background were obtained from Jackson Laboratory (stock #005025). Primers for genotyping: *SMNΔ7*: forward sequence (5’ to 3’) = GATGATTCTGACATTTGGGATG, reverse sequences (5’ to 3’) = TGGCTTATCTGGAGTTTCACAA and GAGTAACAACCCGTCGGATTC (wild-type band: 325bp, mSmn ko: 411bp). Control animals were littermates of mutants as previously reported for the *SMNΔ7* line = *Smn*^+/+^; *SMN2*^+/+^; *SMNΔ7*^+/+^ (Le et al., 2005). C3 compound was dissolved in sterile DMSO (0.4mg/ml in DMSO for the dose of 3mg/kg) and delivered daily via peritoneal injection at a concentration of 3mg/kg from P1 to P30.

Mice from all experimental groups were monitored daily, and the body weight measurements, as well as the righting time and posture time (performed three times and averaged), were recorded as described previously (Mentis et al., 2011). Mice with a 25% reduction of body weight and an inability to right were euthanized to comply with IACUC guidelines for the welfare of experimental animals. Righting time was defined as the time for the pup to turn over on all its four limbs after being placed on its back and maintain its posture for at least 3 seconds. Posture time was defined as the time for which the pup could keep its balance when positioned in its four limbs. The cut-off test time for both tests was 60s. Approximately equal proportions of mice of both sexes were used and aggregated data are presented since gender-specific differences were not found nor have they been previously reported to be affected in SMA.

### Intracellular recordings utilizing a modified *ex vivo* ventral funiculus-ablated mouse spinal cord preparation

The intracellular recordings from motor neurons’ experiments were conducted as previously reported (Mentis et al., 2011; Özyurt et al., 2022; Simon et al., 2017) at P10. The pups were decapitated, eviscerated and the carcass placed in a chamber filled with cold (∼4°C) artificial cerebrospinal fluid (aCSF) to dissect and remove the spinal cord. The aCSF was continuously oxygenated (95%O_2_/5%CO_2_) and contained: 113mM NaCl, 3mM KCl, 1mM NaH_2_PO_4_.H_2_0, 25mM NaHCO_3_, 11mM D-Glucose, 2mM CaCl_2_.H_2_0, and 2mM MgSO_4_.7H_2_0. After the dissection, the L2 – L6 spinal cord segment was glued with the dorsal side facing up and with intact dorsal roots onto an agar block (∼5%) prepared with PBS and 1% FastGreen. An oblique cut (∼45°) was performed at the L2 – L3 region, to visualize the central canal. The agar with the spinal cord was glued into a vibratome chamber (HM 650 V, Microm, Thermo Fisher Scientific, UK) filled with ice-cold (∼4°C) oxygenated aCSF used for the removal of the ventral funiculus (to improve the access to motor neurons). This type of aCSF was different and contained: 130mM K-Gluconate, 15mM KCl, 0.05mM EGTA, 20mM HEPES, 25mM D-Glucose, 3 mM Na-kynurenic acid, 2mM Na-pyruvate, 3mM Myoinositol, 1mM Na-l-ascorbate and the pH was adjusted to 7.4 with NaOH. The vibratome razor blade was aligned with the edge being at the midpoint between the lower end of the central canal and the start of the ventral commissure. The spinal cord was coronally sectioned in a slow pace (0.02 mm/s) to remove the ventral funiculus. The spinal cord was subsequently transferred into a beaker for incubation containing a different aCSF (used for the recording session) at 35°C for 30 minutes. This aCSF was identical to that in which the dissection was performed in and described above. After the 30-minute incubation, the spinal cord was transferred into the recording chamber and perfused with aCSF at room temperature (21 – 23 °C). The dorsal root of the L5 segment was placed into a suction electrode for stimulation.

Motor neurons were visually targeted for patching. At an earlier age (∼P1), pups were anesthetized with isoflurane and the TA muscle was injected with ∼1μl of CTb conjugated with a fluorochrome (CTb-488 or CTb-555). CTb+ labelled motor neurons were selected for recordings with an Eclipse FN1 Nikon microscope outfitted with a digital camera (Moment, Teledyne Photometrics). In some experiments, intracellularly recorded neurons were validated as motor neurons with *post-hoc* immunoreactivity with a ChAT antibody. In these experiments, Neurobiotin was added to the intracellular solution to label the recorded motor neuron *post-hoc*. Patch pipettes were prepared from borosilicate glass (GB200F-10, Science Products Hofheim am Taunus) with a Flaming-Brown puller (P97, Sutter Instruments, CA) to a resistance of ∼3-6 MΩ. The electrodes were then filled with an intracellular solution containing: 150 mM K-D-Gluconate, 10 mM NaCl, 10 mM HEPES, 3 mM Mg-ATP, 0.3mM Na-GTP, 0.05 mM EGTA dissolved in purified water and adjusted to pH 7.3 with KOH and osmolarity of 290–300 mOsm/kg H_2_O (Bornschein et al., 2019). The recorded potentials (DC – 3kHz, Cyberamp, Molecular Devices) were fed to an A/D interface HEKA EPC10/2 amplifier (HEKA humac platform) and amplified with HEKA Patchmaster (HEKA Electronics) at a sampling rate of 10 kHz. In current clamp mode, the membrane passive properties of motor neurons were estimated following brief current injections (300 ms) at 20 pA current steps. Frequency-to-current plots were performed in current clamp mode by injecting long (1sec) current steps (50 pA) of increasing intensity. Excitatory postsynaptic potentials (EPSPs) were recorded from motor neurons in response to a brief (0.2ms) orthodromic stimulation (A365, current stimulus isolator, WPI, Sarasota, FL) of the L5 dorsal root, while holding the cell at membrane potential of −70 mV. The dorsal root was stimulated at 0.1Hz, 1Hz or 10Hz for ten stimuli and the resulting monosynaptic response recorded and analyzed off-line. Data were analyzed offline using HEKA Patchmaster (HEKA Electronics). The monosynaptic component of the EPSP amplitude was measured from the onset of response to 2.5 ms. The latency was determined by measuring the time from the stimulation to the onset of response.

### H-reflex in neonatal and juvenile mice

For neonatal P10 mice, the M-response and H-reflex were recorded *ex vivo* of WT and SMA mutant littermates, as we have reported previously (de Nooij et al., 2015). Animals were decapitated, and the spinal cords were dissected and removed under cold (∼5°C) aCSF containing the following (in mM): 128.35 NaCl, 4 KCl, 0.58 NaH_2_PO_4_-H_2_O, 21 NaHCO_3_, 30D-glucose, 1.5 CaCl_2_-H_2_O, and 1 MgSO_4_-7H_2_O. The sciatic nerve was dissected in continuity with its parent ventral and dorsal roots (L4-L5) to the hindlimb. The spinal cord-hindlimb preparation was then transferred to a customized recording chamber and perfused continuously with oxygenated (95%O_2_/5%CO_2_) aCSF at ∼22°C. A bipolar “hook” electrode was placed on the sciatic nerve for “*en passant*” stimulation. A bipolar concentric needle was inserted into the tibialis anterior (TA) muscle to record EMG activity. The responses of M-response and H-reflex of the TA muscle elicited at incremental stimulation intensities in the sciatic nerve. Of all recordings, the maximum amplitude of M-response and H-reflex were measured. Latencies were measured from the onset of the stimulation artifact to the onset of the M-response and H-reflex.

For juvenile P30 mice, the H-reflex and M-response were recorded *in vivo*. The EMG recordings were performed under isoflurane anesthesia (5% for induction, 1-2% for maintenance, vaporized in O_2_ at a ∼2 liters/min flow rate). A pair of needle electrodes was inserted in the thigh, at the popliteal fossa level, for sciatic nerve stimulation, and another pair was used in the hind paw for plantar muscles EMG recording (one electrode was inserted in the belly of the flexor digitorum brevis (FDB) and the other one in its tendon near the heel). A DC stimulus isolator was used to deliver short pulses (0.1ms) of increasing intensities (0.1µA to 50µA) and at low-frequencies (0.1Hz to 10Hz). The stimulation intensity ranged from sub-threshold level for the Compound Muscle Action Potentials (CMAPs) to twice this intensity (sufficient to elicit maximal M-waves amplitude). The maximal H-reflex amplitude reached throughout this stimulation intensity range was retained and recorded for *post hoc* analysis. Body temperature was monitored and maintained at 38°C with a heating pad throughout the entire recording session and the mouse was euthanized immediately thereafter with an intraperitoneal injection of tribromoethanol (300mg/kg) followed by cervical dislocation.

### Immunohistochemistry on murine and human spinal cord tissue

The detailed protocols used for immunohistochemistry in mouse tissues have been previously reported (Buettner et al., 2022; Buettner *et al*., 2021). Briefly, mice were perfused with 1x PBS and 4% PFA following 4% PFA post-fixation overnight at 4°C. The following day, the spinal cords were removed and the lumbar L5 segments were identified by the ventral roots and briefly washed with PBS. Subsequently, single segments were embedded in warm 5% agar and serial transverse sections (75μm) were cut at the vibratome. The sections were blocked with 5% normal donkey serum in 0.01M PBS with 0.3% Triton X-100 (PBS-T; pH 7.4) for 90 minutes and incubated overnight at room temperature in different combinations of the primary antibodies (Table 1: List of primary antibodies). VGluT1 antibodies were used as a marker for proprioceptive synapses, ChAT antibodies were used to label motor neurons. The following day, after 6 washes of 10 minutes with PBS, secondary antibody incubations were performed for 3 hours with the appropriate species-specific antiserum coupled to Alexa488, Cy3 or Alexa647 (Jackson labs) diluted at 1:1000 in PBS-T. After secondary antibody incubations, the sections were washed 6 times for 10 minutes in PBS, mounted on slides, and cover-slipped with an anti-fading solution made of Glycerol:PBS (3:7) (Simon et al., 2017).

**Table 1:**
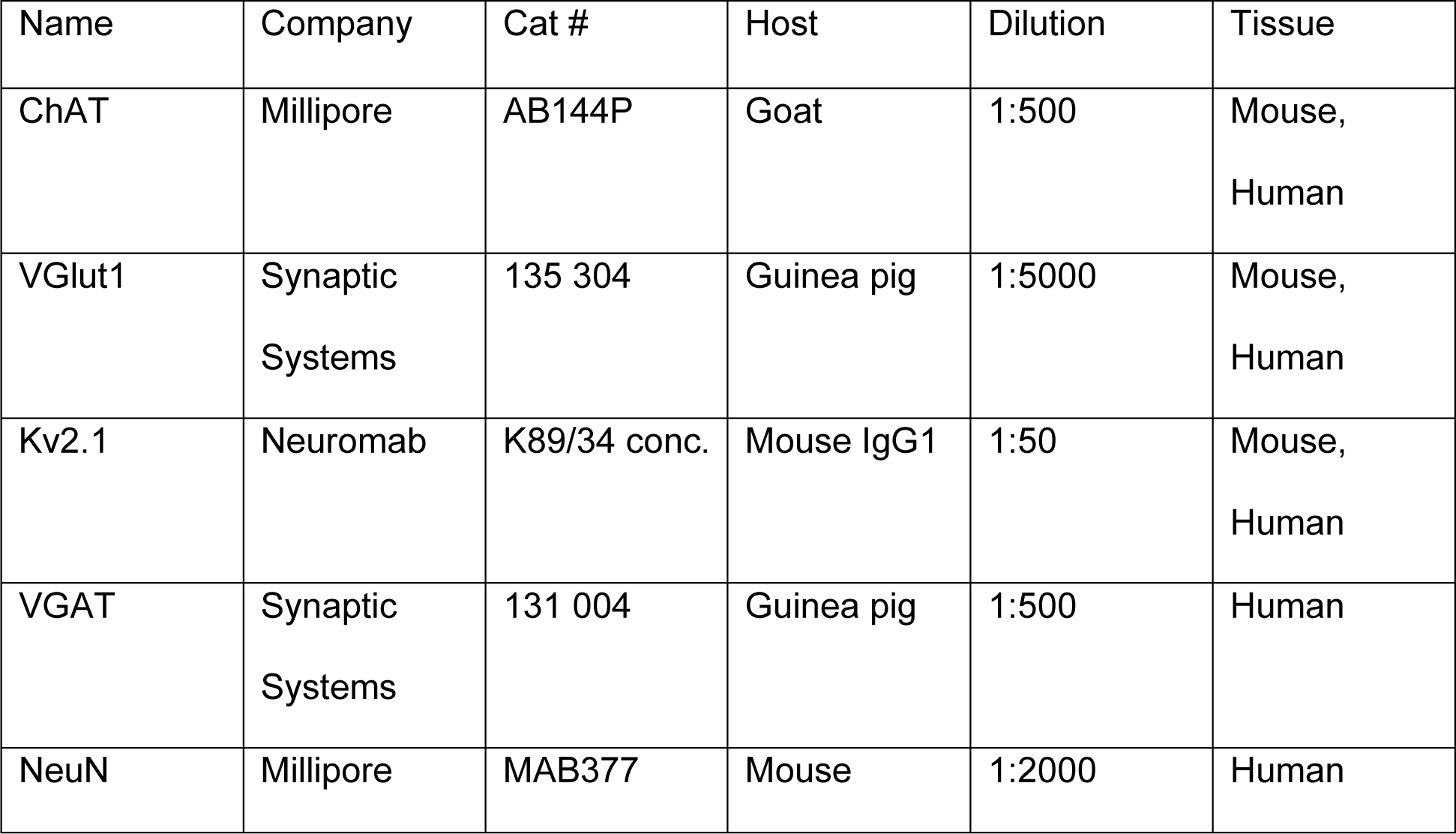
List of primary antibodies.

For immunohistochemistry on postmortem human tissue, some of the thoracic and lumbar segments of the spinal cord from patients with SMA and non-SMA controls were collected at Johns Hopkins University School of Medicine, Baltimore, MD, while some others were collected from Children’s National Hospital, George Washington University, School of Medicine and Health Sciences, in Washington, DC (IRB:15350) or at Columbia University in NY, NY. Expedited autopsies were conducted under parental- or patient-informed consent in strict observance of the legal and institutional ethical regulations. Tissues from patients with SMA and age-matched controls with a short postmortem interval were included. All patient and control tissue used in this study is listed in Table 2. Human spinal cord tissue was cryoprotected with a 15% sucrose solution for at least 2h until the tissue sank to the bottom of the tube. Afterwards, the tissue was transferred to a 30% sucrose solution and stored at 4°C overnight. The next day, the spinal cord tissue was embedded in Sakura Tissue Tek O.C.T. Compound and frozen in 2-Methylbutan cooled with liquid nitrogen. Two spinal cords (one SMA and one control) were flash frozen immediately upon removal from the patient and without fixation. Tissues were cut at a Leica CM3050 S cryostat into 20µm serial transverse sections at −20°C and subsequently stored at −80°C. Human spinal cord sections were incubated for 20min in Polyscience L.A.B. solution for antigen retrieval at room temperature. Afterwards they were washed 3x with PBS and blocked in 5 % normal donkey serum in 0.03% PBS-T for 90min. They were then incubated with the appropriate primary antibodies (see Table 1) at 4°C overnight followed by 3x 10min washes with PBS and incubated for 3h with the appropriate secondary antibodies diluted 1:1000 in PBS-Triton. VGAT was used as a marker for inhibitory synapses and NeuN in combination with ChAT to identify α-MNs. After washing 3x 10 minutes with PBS the slides were cover-slipped with Glycerol: PBS (3:7) medium. In a few experiments, triplicates of analysis could not be performed due to over-fixation or reduced availability of human tissue.

**Table 2:**
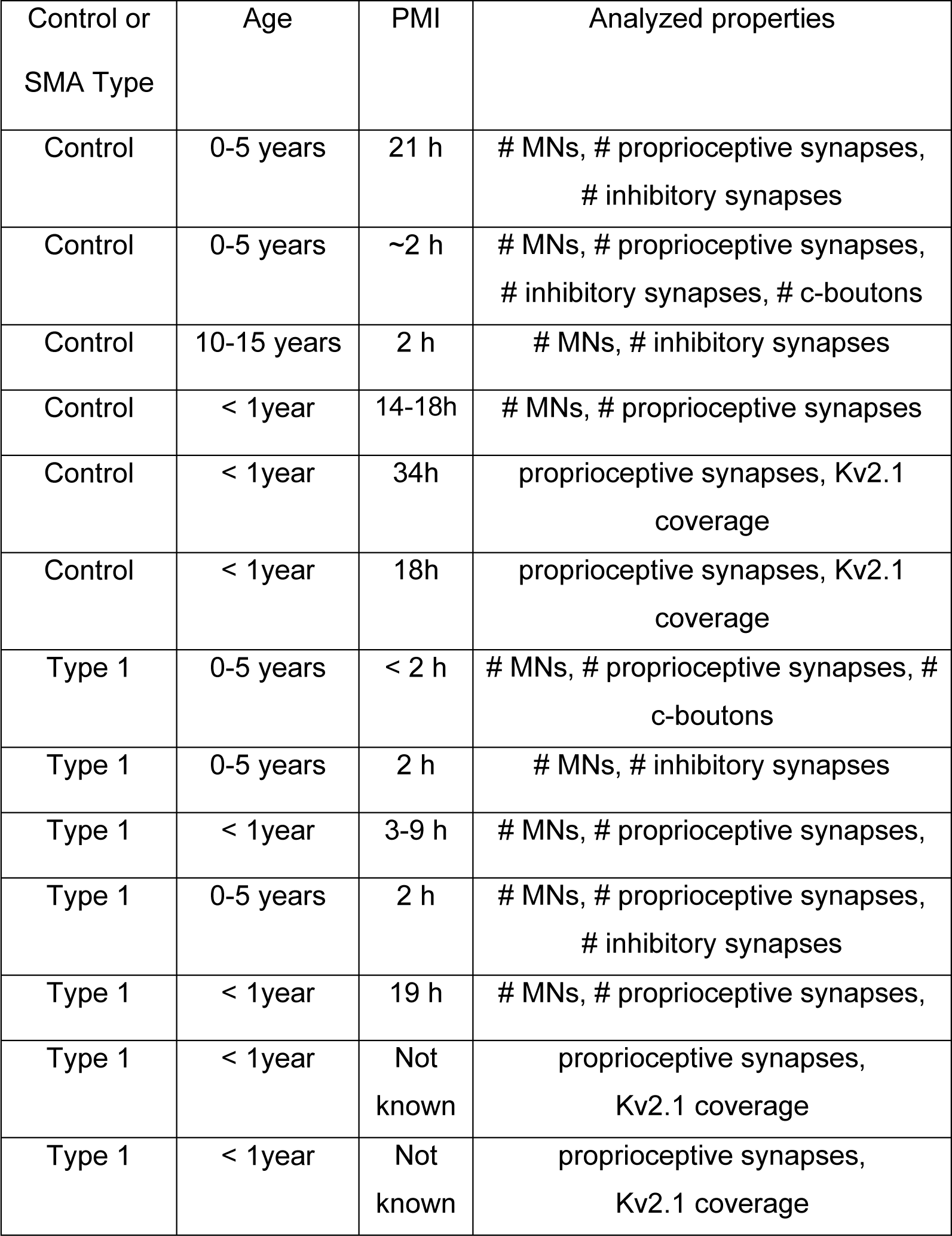
List of patient tissue used for immunohistochemical analysis.

**Table 3:**
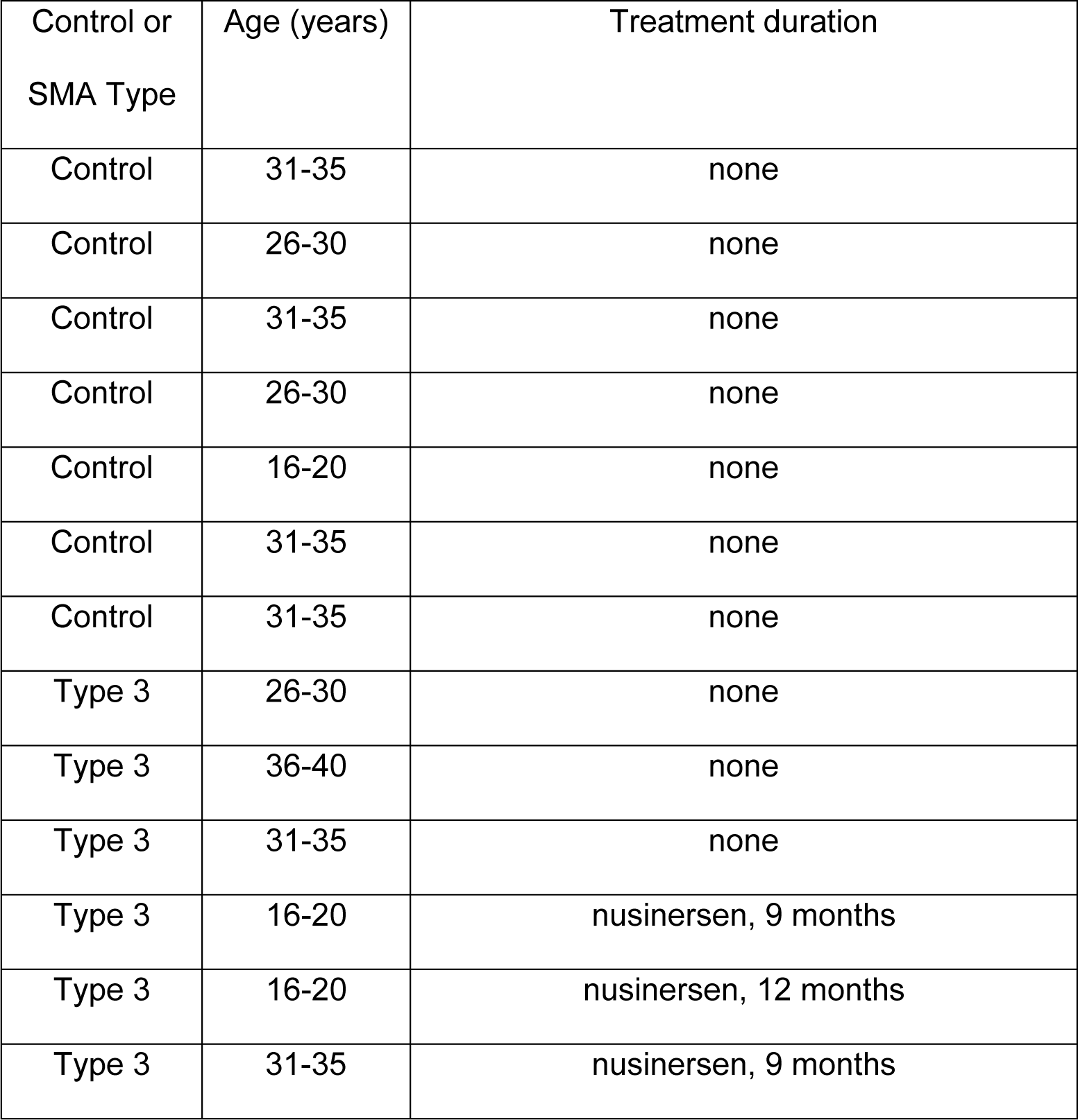
List of patients for EMG studies.

### Confocal microscopy and analysis

Spinal cord sections from mice and humans (see Table 2: List of patient tissue used for immunohistochemical analysis) were imaged using SP8 Leica confocal microscopes or an Olympus FV300 inverted microscope. Sections for motor neuron counting were scanned using a 20x objective. Motor neurons were counted off-line from z-stack images (collected at 4μm intervals in the z-axis) from an entire spinal cord segment for mice or per section for human motor neurons. At least three sections were analyzed for human motor neuron counts and NeuN was used to distinguish between alpha (NeuN+) and gamma (NeuN-) motor neurons. Only ChAT+ motor neurons located within the ventral horn containing the nucleus were counted to avoid double counting from adjoining sections.

Quantitative analysis of VGluT1 (proprioceptive synapses), VGAT (inhibitory synapses) and C-boutons (appearing as intense ChAT+ synapses around the motor neuron cell membrane) on motor neurons was performed on image stacks of optical sections scanned using a 40x oil or 63x glycerol objective throughout the whole section thickness at 0.4μm z-steps. The number of VGluT1+ and ChAT+ synapses were counted over the entire surface of the murine motor neuron soma as well as on primary dendrites for VGluT1+ synapses up to a distance of 50µm from the soma using Leica LASX software as previously described (Buettner et al., 2022). VGAT synapses were quantified as the number of synapses per perimeter of the motor neuron soma. We analyzed three different optical planes for each motor neuron to ensure consistency. First, we scanned the optical plane in the center of the motor neuron defined by the largest area of its soma and a visible nucleus. From that point, we scanned two additional optical planes with a distance of 6µm above and below from the first plane. Then we quantified the number of VGAT+ synapses onto ChAT+ motor neurons and the perimeter of the soma on each plane by Leica LASX software. Division of synaptic number by perimeter resulted in synaptic density of each panel. We averaged the synaptic density of all three panels for each individual motor neuron. At least 10 motor neurons per mouse or human were quantified as previously described (Buettner et al., 2021).

Kv2.1 coverage was quantified as previously described (Fletcher et al., 2017). Briefly, single optical plane high resolution images were taken with a 63x oil immersive objective. Only motor neurons with their nucleus present in the optical plane were analyzed. Using the FIJI Image J software, a line was drawn around the perimeter of the motor neuron, excluding any dendrites, to acquire the fluorescent signal intensity for Kv2.1. To measure a baseline of Kv2.1 intensity, a line was drawn inside the soma, avoiding the nucleus. Only Kv2.1 intensity signals 3x standard deviations above the baseline intensity were considered as Kv2.1 expression and their coverage along the cell perimeter was calculated as a percentage.

### H-Reflex in SMA and control patients

Healthy and SMA individuals were seen at the Department of Neurology at Columbia University Irving Medical Center. All individuals consented to H-reflex testing. The individuals were prone positioned and reclined with the head and arms supported to reduce the variability of the H-reflex and to preserve the individual’s comfort. We used silver/silver chloride 20mm diameter round surface electrodes for recording and placing the active electrode over the soleus muscle. The reference electrode was placed on the Achilles tendon, and the ground electrode was placed between the stimulation and recording sites. The tibial nerve was stimulated by a surface probe, positioning the cathode over the tibial nerve at the popliteal fossa and the anode side more distal. A 1 ms duration pulse was delivered with increasing intensity. To reduce the effects of post-activation depression, we waited at least 10secs between stimuli. The maximal H-reflex was determined once higher stimulation intensities produced a smaller response, typical of this reflex. The maximal H-reflex amplitude was used to calculate the H/M ratio. The EMG equipment used was the Natus Viking EDX system (version 22). The disposable electrodes used were manufactured by Natus Medical, Inc.

### Proprioception test in control and Type 3 SMA participants

The experimental protocols for this test were approved by the University of Pittsburgh Institutional Review Board (IRB) (protocol STUDY21080158) under an abbreviated investigational device exemption. Three male individuals with Type 3 SMA and five controls took part in this test. Prior to their involvement, participants underwent an informed consent process, in accordance with the procedure approved by the IRB. SMA participants were part of a clinical trial publicly available on ClinicalTrials.gov (NCT05430113) on epidural electrical stimulation.

All SMA participants were able to stand and walk (with the assistance of walking aids for SMA02 and SMA03). SMA01 had mild motor deficits with an HFMSE score at enrollment of 60/66 points. He was not under any treatment during the test. SMA02 had the most severe motor deficit with an HFMSE score of 38/66 points. He was being treated with nusinersen. SMA03 had a moderate motor deficit with an HFMSE score of 49/66 points. During the test he was being treated with nusinersen. The Hammersmith Functional Motor Scale Expanded (HFMSE) was developed for patients with SMA who are ambulatory. The HFMSE consists of 33 items that are scored either 0, 1 or 2. A score of 2 is assigned to participants who achieve the motor task without any compensatory strategies. Attempted movements or items achieved with compensation are scored a 1. A score of zero is assigned to those unable to perform the task.

The test for proprioception was performed using the Humac Norm Cybex system (Computer Sports Medicine Inc., Stoughton, US). The constant angular velocity was 0.5° s-1. During the experiment, participants’ visual and aural information were occluded using foam blindfolds and headphones with white noise. At the beginning of each trial, the knee joint is positioned at 45° degrees of extension. The participant is informed of the beginning of the trial by a tap on the shoulder. The trial starts after a randomized time delay (0-10s) to evaluate false positive detections. At a constant angular velocity, the platform passively moves the limb, eliciting flexion, or extension of the knee. As soon as the direction of the movement is detected, the participant presses a button to stop the platform, and the perceived direction is verbally reported. The detection angle (the difference between the final and initial angle displacement) as well as the error rate (number of times the participant reported wrong directions) are then calculated.

### 6-minute-walk test (6MWT) distance and fatigability in SMA patients

The 6-minute walk test (6MWT) was performed at the Department of Rehabilitation and Regenerative Medicine at Columbia University Irving Medical Center. Distance walked in meters was measured and fatigability was calculated. All participants signed informed consent approved by the Columbia University’s Irving Medical Center Institutional Review Board (AAAE8252). The 6MWT is an objective evaluation of exercise capacity and is representative of a person’s ability because the test intensity is self-selected (Solway et al., 2001). The 6MWT has been used to assess function (Andersson et al., 2006; Takeuchi et al., 2008) and has been accepted by regulatory agencies as a clinically meaningful endpoint in other neurologic disorders. It has been described and studied extensively in SMA as we (Montes et al., 2019; Montes et al., 2010) and others (Pera et al., 2017) have previously reported. Briefly, SMA patients were instructed to walk as fast as possible along a 25-meter line marked course on a linoleum floor, turn around a marker cone, and then return in the opposite direction and repeat this loop as often as possible for 6 minutes. The course had a start line, placed horizontally at the beginning, with smaller horizontal lines placed every 1 m. The participants wore rubber-soled shoes appropriate for walking. Participants were allowed to rest without sitting, if necessary. Fatigability was calculated by subtracting the distance walked in the sixth minute from the distance walked in the first minute. Measurements are expressed as percentages, where a positive value represents fatigue, as previously reported (Montes et al., 2013).

### Statistics

Results are expressed as mean ± standard deviation (SD) from at least three independent experiments using three or more animals/patients per experimental group unless differently indicated in the figure legend. Shapiro-Wilk normality test was used to test for symmetric data distribution. For comparison of two groups: For a parametric distribution of two groups an unpaired *t* test or multiple *t* test was used when standard deviation (SD) of both populations were the same. When SDs were not equal, a Welch’s *t* test was used. Mann-Whitney or multiple Mann-Whitney test was used for nonparametric distribution. For comparison of three groups: A one-way ANOVA with Tukey multiple comparison tests was applied for a parametric distribution of three groups. For a nonparametric distribution of three groups a Kruskal-Wallis test with Dunn’s correction was applied. When SD were not equal, a Welch’s ANOVA with Dunnett’s T3 multiple comparisons test was applied. For time course comparison of more than two groups a two-way ANOVA with Tukey multiple comparison test was applied. N values indicate either cells or animals/patients as indicated in the figure legend. GraphPad Prism 10 was used for all statistical analyses and p values are indicated in the figure.

## Data Availability

All data produced in the present study are available upon reasonable request to the authors

## SUPPLEMENTAL FIGURE LEGENDS

**Supplemental Figure 1.**
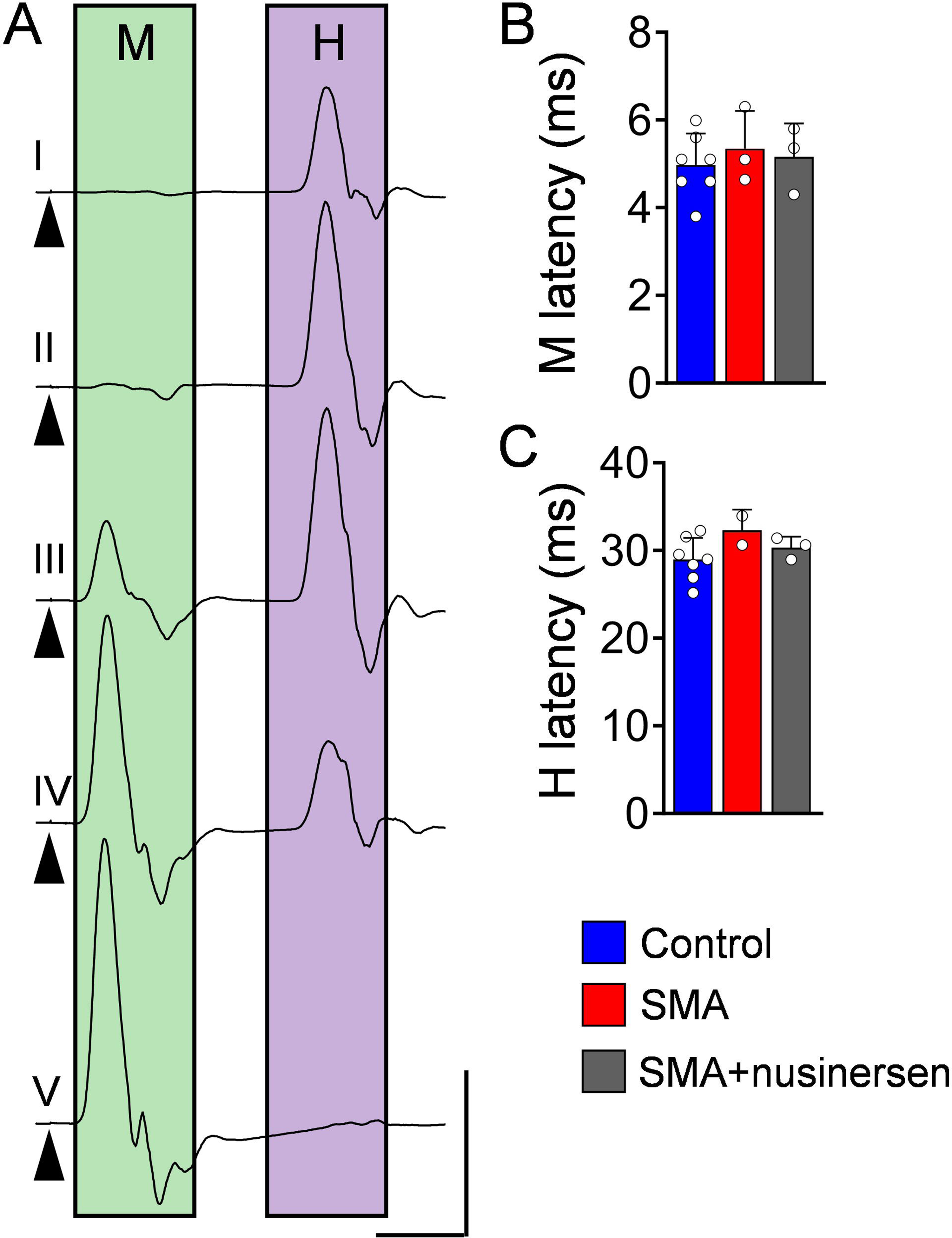
(associated with Fig.2). Latencies of the M-response and H-reflex are unaltered in SMA patients. (**A**) Soleus EMG recordings following incremental stimulation intensities (from top to bottom) of the tibial nerve in a control subject. Note: highest response of H in III and M in V. Arrowheads indicate stimulus artefacts. Scale bar: 5mV, 10ms. (**B**) latency of M-response and (**C**) latency of H-reflex for control (N = 7) and SMA-untreated (N = 3) and nusinersen-treated (N = 3) Type 3 SMA patients. Data represent means and SD. Statistical analysis was performed using one-way ANOVA with Tukey multiple comparison test (B, C).

**Supplemental Figure 2.**
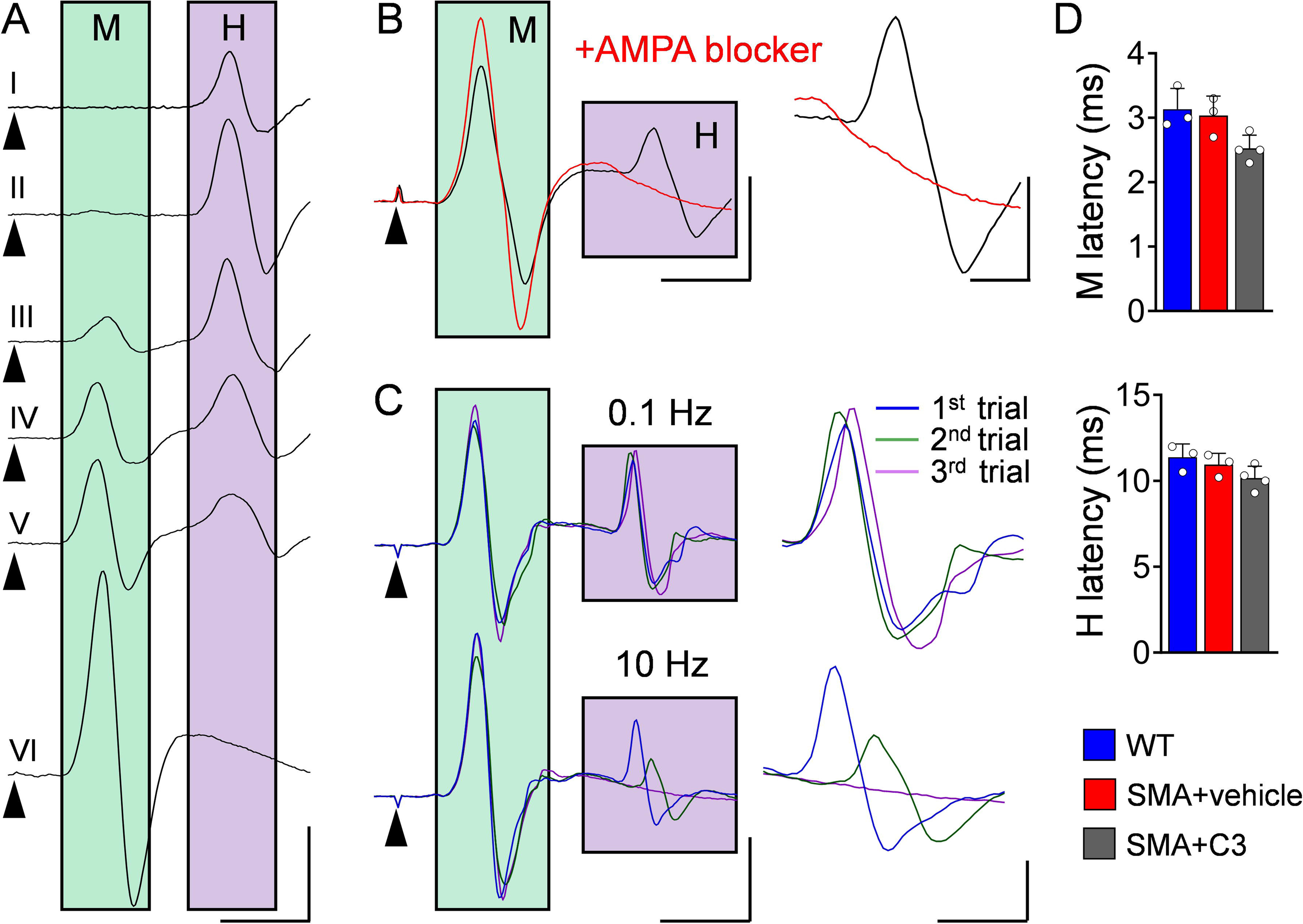
(associated with Fig.3). Latencies of M- and H-response are unaltered in neonatal SMA mice. (**A**) EMG recordings from TA muscle following incremental stimulation intensities (from top to bottom) of the sciatic nerve in a P10 WT mouse. Note: highest response of H in II and M in VI. Arrowheads indicate stimulus artefacts. Scale bar: 0.5mV, 5ms. (**B**) EMG recordings from TA muscle following stimulation of the sciatic nerve from a P10 WT mouse before (black trace) and after the AMPA blocker NBQX (red) application. Scale bar: 0.5mV, 5ms. Right traces show magnification of H-response. Scale bar: 0.2mV, 2ms. (**C**) EMG recordings from a TA muscle following 0.1Hz (upper panel) and 10 Hz (lower panel) repetitive stimulation of the sciatic nerve from a P10 WT mouse. Scale bar: 0.5mV, 5ms. Right traces show magnification of H-response. Scale bar: 0.2mV, 2ms. (**D**) latency of the M-response (top) and H-reflex (bottom) from WT (N = 3), SMA+vehicle (N = 3) and SMA+C3 (N = 4) using *ex vivo* spinal cords from mice at P10. Data represent means and SD. Statistical analysis was performed using an one-way ANOVA with Tukey multiple comparison test (D).

**Supplemental Figure 3.**
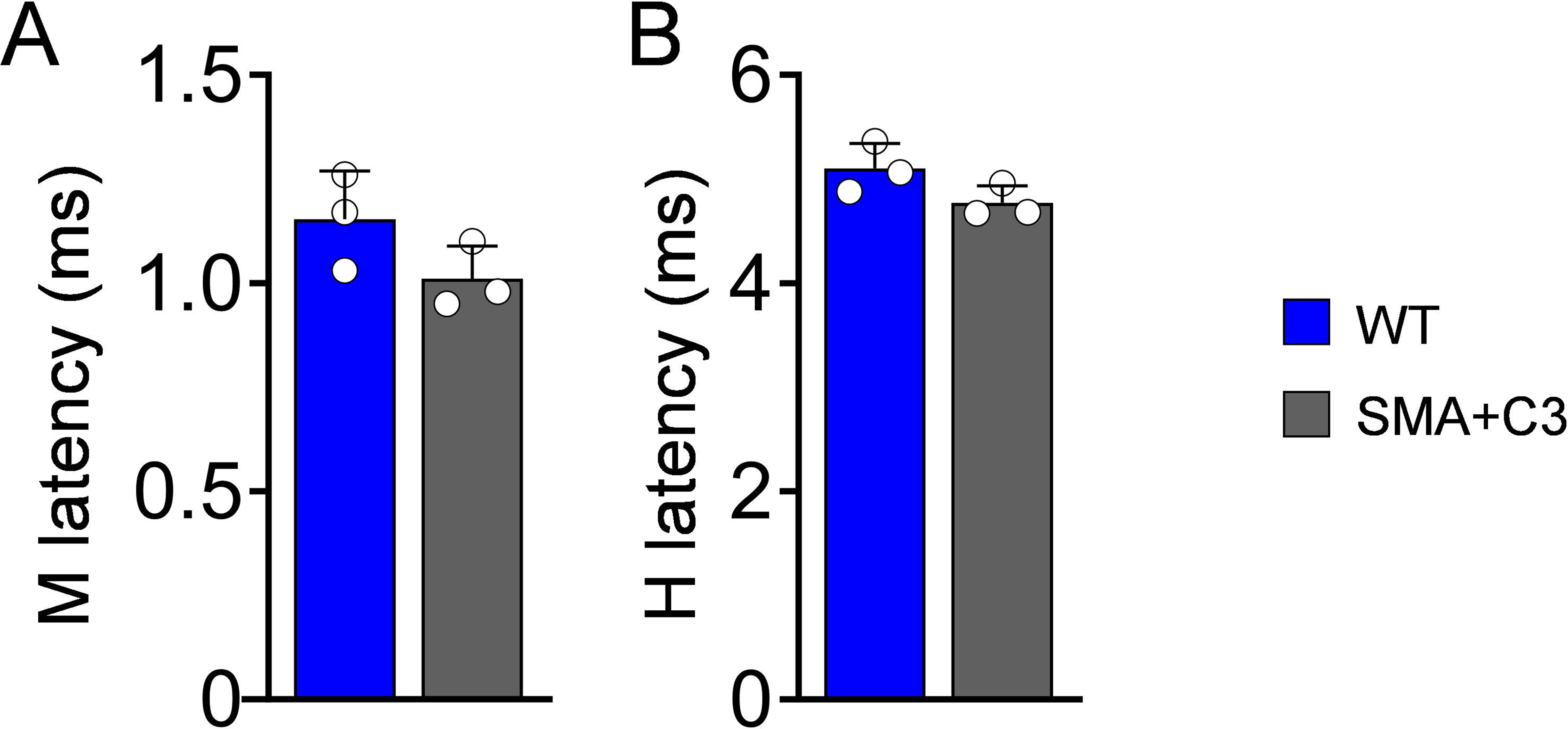
(associated with Fig.4). Latencies of M- and H-responses are unaltered in juvenile SMA mice. The latency for M-response (**A**) and H-reflex (**B**) in WT (N = 3) and SMA+C3 (N = 3) mice at P30. Data represent means and SD. Statistical analysis was performed using an one-way ANOVA with Kruskal-Wallis test with Dunn’s correction.

**Supplemental Figure 4.**
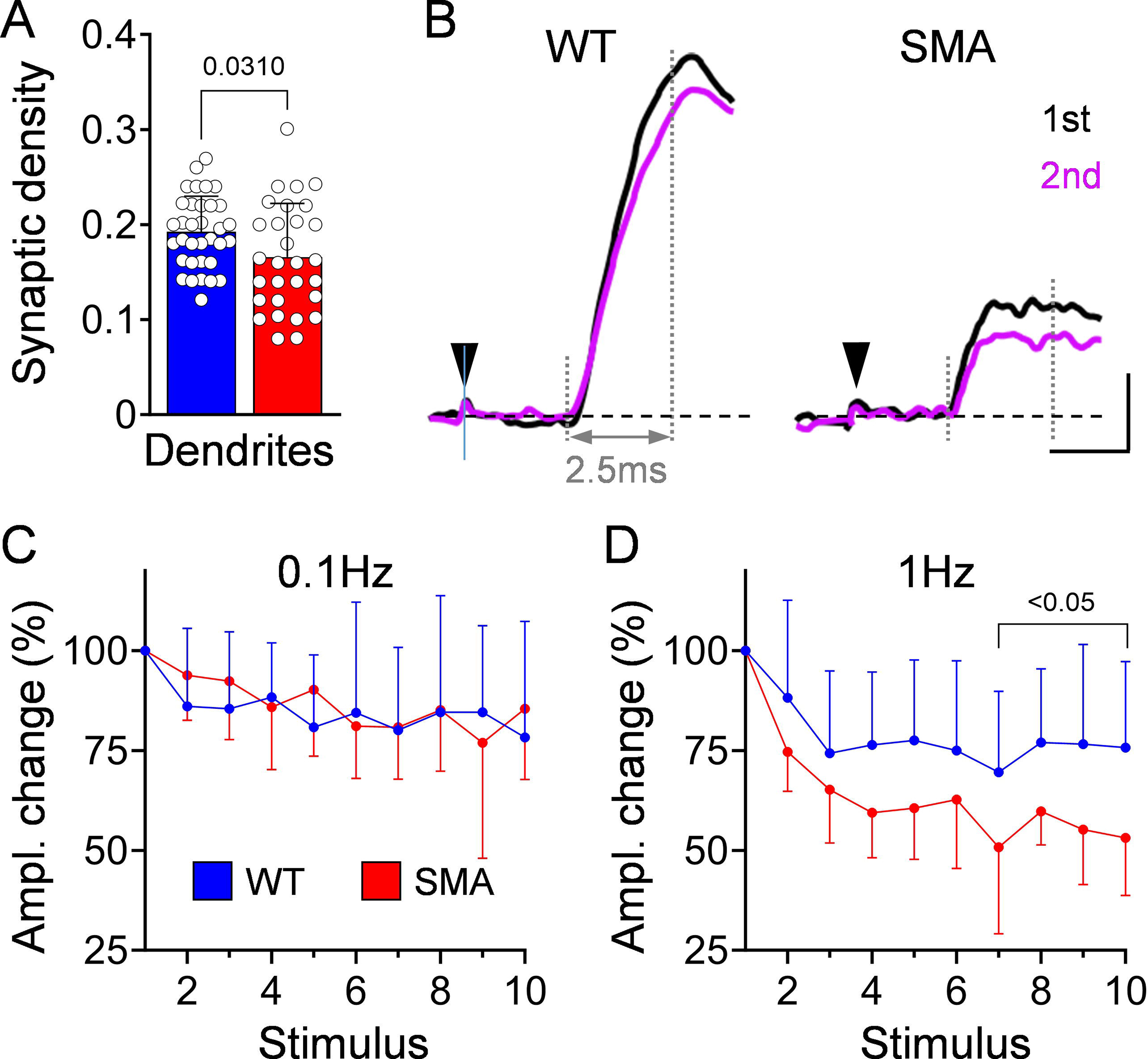
(associated with Fig.5). The number of proprioceptive synapses are mildly affected on dendrites of SMA motor neuron innervating distal muscles and reduction of their EPSPs at different frequencies. (**A**) Density of proprioceptive synapses on proximal dendrites of motor neurons (0-50µm from soma) for WT (*n* = 34, N = 4 mice) and SMA (*n* = 29, N = 3 mice) at P10. (**B**) First (black) and second (magenta) EPSP responses elicited in motor neurons after 1Hz dorsal root stimulation in WT and SMA mice at P10. Arrowheads indicate stimulus artefact. Vertical dotted line marks peak EPSP amplitude measured at 2.5ms after the onset of response. Scale bar: 2mV, 2ms. Changes of EPSP amplitude following 10 stimuli at (**C**) 0.1Hz and (**D**) 1Hz normalized to the first response for WT (*n* = 10) and SMA (*n* = 9) motor neurons at P10. Data represent means and SD. Statistical analysis was performed using unpaired *t* test for (A) and multiple Mann-Whitney test (C, D).

**Supplemental Figure 5.**
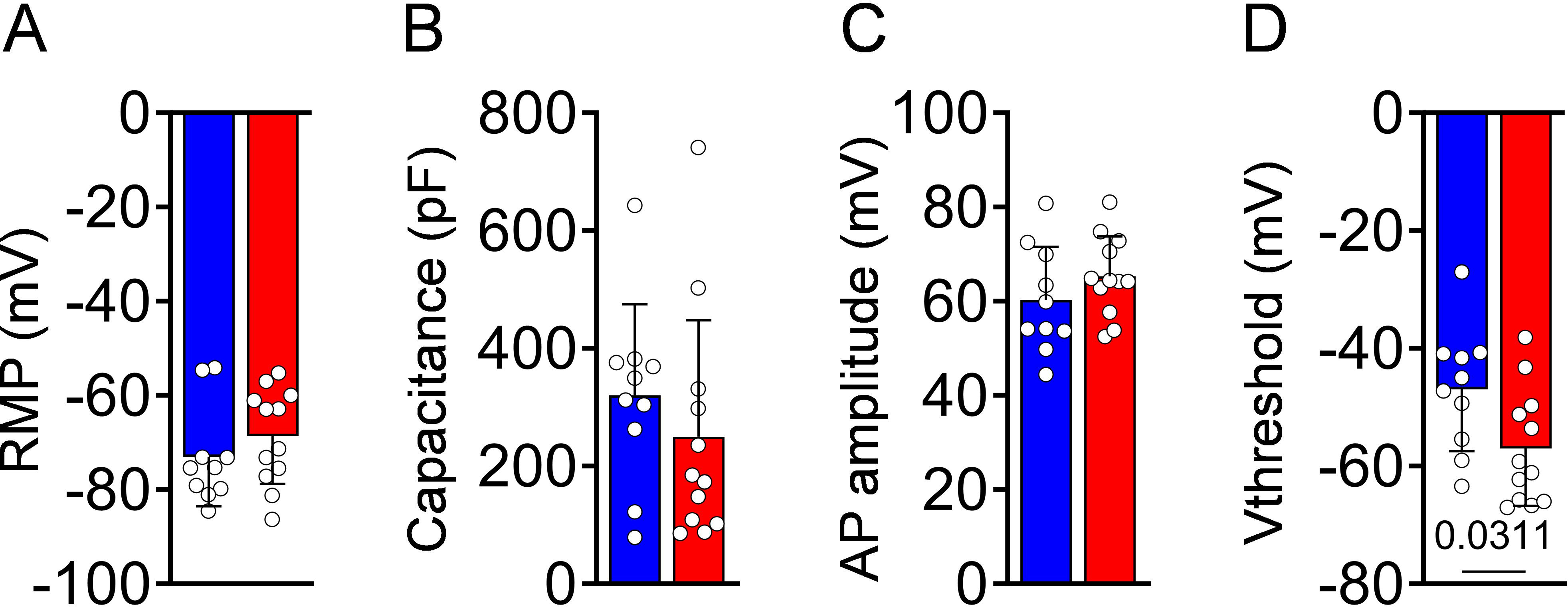
(associated with Fig.6). Motor neurons innervating distal muscles have a lower firing threshold in SMA mice. (**A**) Resting membrane potential (RMP), (**B**) Capacitance, (**C**) Action potential amplitude and (**D**) V_threshold_ in WT (*n* = 10) and SMA (*n* = 12) motor neurons at P10. Data represent means and SD. Statistical analysis was performed using Mann-Whitney test for (A) and unpaired *t* test for (B-D).

**Supplemental Figure 6.**
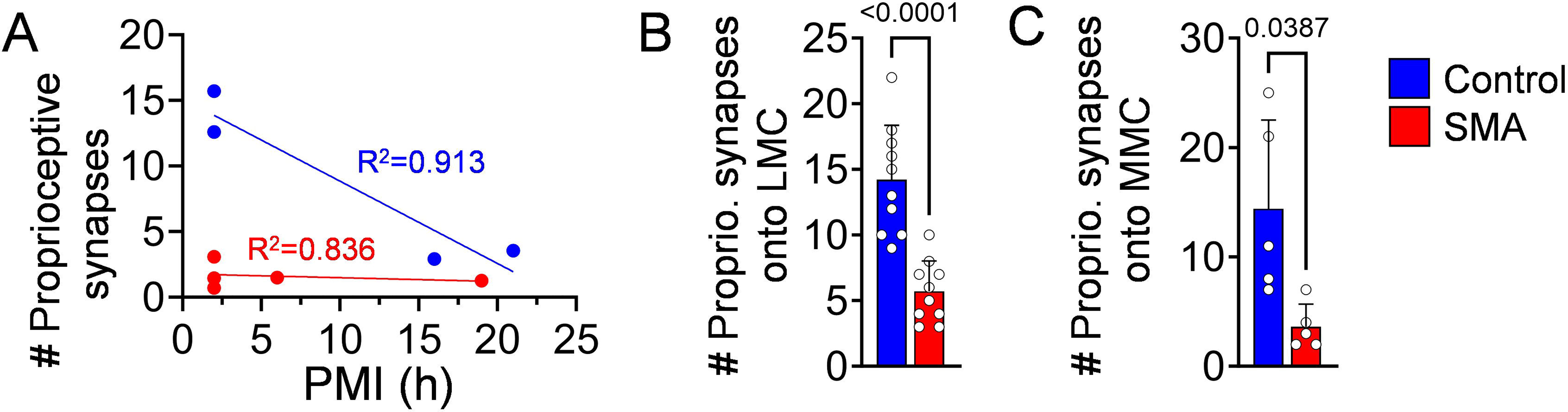
(associated with Fig.7). Reduction of proprioceptive synapses on spinal motor neurons and relation with PMI. (**A**) Relationship of proprioceptive synapses and post-mortem interval (PMI) from control (N = 4) and Type I SMA (N = 5) spinal cords. Number of proprioceptive synapses (**B**) lumbar lateral motor neurons (LMNs) from control (*n* = 10 MNs, N = 1 subject) and SMA type I (*n* = 10 MNs, N = 1 patient) patients and (**C**) lumbar medial motor neurons (MMNs) from control (*n* = 5 MNs, N = 1 subject) and SMA type I patients (*n* = 5 MNs, N = 1 patient). Data represent means and SD. Statistical analysis was performed using a simple linear regression (A), Welch’s *t* test for (B,C).

## ACKNOWLEDGMENTS

We thank the generous autopsy tissue donations from patients and their families and the financial support provided by the SMA Foundation. We also want to thank all SMA patients and healthy participants in the H-reflex investigations. We thank Dr Michio Hirano for providing critical comments to our study and Dr Marco Beato and Dr Filipe Nascimento for providing guidance to establish the ventral funiculus-ablated *ex vivo* spinal cord preparation.

This work is supported by R01-NS078375 (GZM), R01-NS125362 (GZM), R01-AA027079 (GZM), The SMA Foundation (GZM), Project ALS (GZM); German Research Foundation grants SI-1969/2-1 (CMS), SI-1969/3-1 (CMS), SMA-Europe (CMS), CMS was also the recipient of Young Investigator award by ROCHE; R01-NS102451 (LP), R01-NS114218 (LP), R01-NS116400 (LP); SMA Foundation (CJS), R35 NS122306 (NINDS to CJS); K12HD001399 (PK), American SIDS Institute (PK), Raynor Cerebellum Project (PK); K01-HD084690 (JM) and Cure SMA (CU18-2886; JM), Grant from F. Hoffmann–La Roche (MC).

## AUTHOR CONTRIBUTIONS

CMS and GZM conceived the study; CMS, ND, JM, FG, EC, LS, JMB, JGP, GP-O, SE, SD, JLG, performed experiments; CMS, ND, JM, FG, EC, LS, JMB, JGP, GP-O, SE, SD, JLG, LHW, EP, MC, LP, DCD, GZM analyzed the data; JM, LHW, DCD performed clinical evaluations; PK, WKC, CJS provided postmortem tissue; CMS, LP and GZM wrote the paper with contributions from all authors.

